# Determinants of SARS-CoV-2 transmission to guide vaccination strategy in an urban area

**DOI:** 10.1101/2020.12.15.20248130

**Authors:** Sarah C. Brüningk, Juliane Klatt, Madlen Stange, Alfredo Mari, Myrta Brunner, Tim-Christoph Roloff, Helena M.B. Seth-Smith, Michael Schweitzer, Karoline Leuzinger, Kirstine K. Søgaard, Diana Albertos Torres, Alexander Gensch, Ann-Kathrin Schlotterbeck, Christian H. Nickel, Nicole Ritz, Ulrich Heininger, Julia Bielicki, Katharina Rentsch, Simon Fuchs, Roland Bingisser, Martin Siegemund, Hans Pargger, Diana Ciardo, Olivier Dubuis, Andreas Buser, Sarah Tschudin-Sutter, Manuel Battegay, Rita Schneider-Sliwa, Karsten M. Borgwardt, Hans H. Hirsch, Adrian Egli

## Abstract

**Background:** Transmission chains within small urban areas (accommodating∼30% of the European population) greatly contribute to case burden and economic impact during the ongoing COVID-19 pandemic, and should be a focus for preventive measures to achieve containment. Here, at very high spatio-temporal resolution, we analysed determinants of SARS-CoV-2 transmission in a European urban area, Basel-City (Switzerland). Methodology. We combined detailed epidemiological, intra-city mobility, and socioeconomic data-sets with whole-genome-sequencing during the first SARS-CoV-2 wave. For this, we succeeded in sequencing 44% of all reported cases from Basel-City and performed phylogenetic clustering and compartmental modelling based on the dominating viral variant (B.1-C15324T; 60% of cases) to identify drivers and patterns of transmission. Based on these results we simulated vaccination scenarios and corresponding healthcare-system burden (intensive-care-unit occupancy). Principal Findings. Transmissions were driven by socioeconomically weaker and highly mobile population groups with mostly cryptic transmissions, whereas amongst more senior population transmission was clustered. Simulated vaccination scenarios assuming 60-90% transmission reduction, and 70-90% reduction of severe cases showed that prioritizing mobile, socioeconomically weaker populations for vaccination would effectively reduce case numbers. However, long-term intensive-care-unit occupation would also be effectively reduced if senior population groups were prioritized, provided there were no changes in testing and prevention strategies. Conclusions. Reducing SARS-CoV-2 transmission through vaccination strongly depends on the efficacy of the deployed vaccine. A combined strategy of protecting risk groups by extensive testing coupled with vaccination of the drivers of transmission (i.e. highly mobile groups) would be most effective at reducing the spread of SARS-CoV-2 within an urban area.

**Author summary:** We examined SARS-CoV-2 transmission patterns within a European city (Basel, Switzerland) to infer drivers of the transmission during the first wave in spring 2020. The combination of diverse data (serological, genomic, transportation, socioeconomic) allowed us to combine phylogenetic analysis with mathematical modelling on related cases that were mapped to a residential address. As a result we could evaluate population groups driving SARS-CoV-2 transmission and quantify their effect on the transmission dynamics. We found traceable transmission chains in wealthier or more senior population groups and cryptic transmissions in the mobile, young or socioeconomic weaker population groups - these were identified as transmission drivers of the first wave. Based on this insight, we simulated vaccination scenarios for various vaccine efficacies to reflect different approaches undertaken to handle the epidemic. We conclude that vaccination of the mobile inherently younger population group would be most effective to handle following waves.

## Introduction

Efforts to understand transmission of SARS-CoV-2 have been undertaken at different scales including at a global level^1–3^, across continents (Europe and North America^4^), within countries (Austria^5^, Brazil^6^, France^7^, Iceland^8^, South Africa^9^, Thailand^10^) and in large cities (Beijing^11^, Boston^12^, Houston^13^, and New York City^14–16^). In Europe, ∼30% of the population live in small urban areas (10*k* − 300*k* inhabitants)^17^, which accordingly play a major role in SARS-CoV-2 transmission yet have not been studied. Moreover, to date city-based studies of SARS-CoV-2 transmission have very limited resolution in terms of the proportion of sequenced positive cases (incomplete transmission chains), have a paucity of socioeconomic or mobility data (incomplete determinants), or fail to combine analysis of transmission clusters with quantitative, descriptive models accounting for population mixing^18^. Of those studies describing the distribution of cases together with changes in mobility, none to date rigorously study socio-economic differences between city quarters as determinants of transmission^13,15,16,19^. An integrated model considering all of these factors (including epidemiological, geographic, mobility, socioeconomic, transmission dynamics information) is anticipated to provide profound insights into the determinants underpinning transmission, that can be used to guide the delivery of vaccines. We here present such an integrated analysis for Basel-City which is part of a metropolitan area, a functional urban area, and a European cross-border area as outlined in the supplement. Basel-City is hence representative of other areas in the EU classified as such. Local interventions are most effective in cutting transmission chains in families, and small community networks^20,21^ that represent well-defined (phylogenetic or epidemiological) clusters. However, most infections are acquired from unknown sources and transmitted cryptically making it essential to identify the key determinants and transmission routes at the city-level to improve interventions and vaccination campaigns. To address this challenge, we have combined phylogenetic cluster analysis^22^ and compartmental ordinary differential equation (ODE) modelling based on high density (81% of reported cases assessed), and high resolution (spatial:housing blocks, temporal: day-by-day) epidemiological, mobility, socioeconomic, and serology (estimate unreported cases) data-sets from the first COVID-19 wave (February-April 2020). Whole genome sequencing (WGS) of all included cases (44% of all cases successful) allowed the analysis be restricted to a single, dominant viral variant (B.1-C15324T, 60% of cases ^23^). This ensured that our analyses focused on inherently related cases and enabled estimation of effective reproductive numbers for different socioeconomic and demographic population subgroups to provide the basis for vaccination scenario building.

## Results

### SARS-CoV-2 spread and clustering

We observed 29 viral lineages in Basel-City (Figure S8), with 247 genomes (60.0%) belonging to the B.1-C15324T variant^23^ (Figure 2A, B). Applying a genetic divergence threshold, a total of 128 phylogenetic clusters were determined across all samples, of which 70 belonged to lineage B.1-C15324T (Figure 2C). Mapping phylogenetic clusters onto tertiles of socioeconomic and demographic determinants, we found that for median income, T1 contained the most and T3 the least clusters (Figure 2C). Most within- and among-tertile transmission clusters were spread randomly, except for significant within-tertile transmissions among high median income households (T3) (Figure 2D). Further, we observed that SARS-CoV-2 isolates were more likely to belong to the same phylogenetic cluster for blocks with either the highest living space per person, lowest share of 1-person households, or highest seniority (Figure 2E). This is also true among people living in the more affluent quarters Riehen, Bruderholz, Am Ring, and Iselin, who transmitted the virus in their social networks either in the same quarter or in the same socioeconomic rank (Figure S7). By contrast, positive cases that belong to lower socioeconomic/demographic tertiles (either lower income, less living space, or younger age) are less likely to be members of the same tight phylogenetic cluster, indicating cryptic transmission predominates among lower socioeconomic and younger demographic groups.

**Figure 1:**
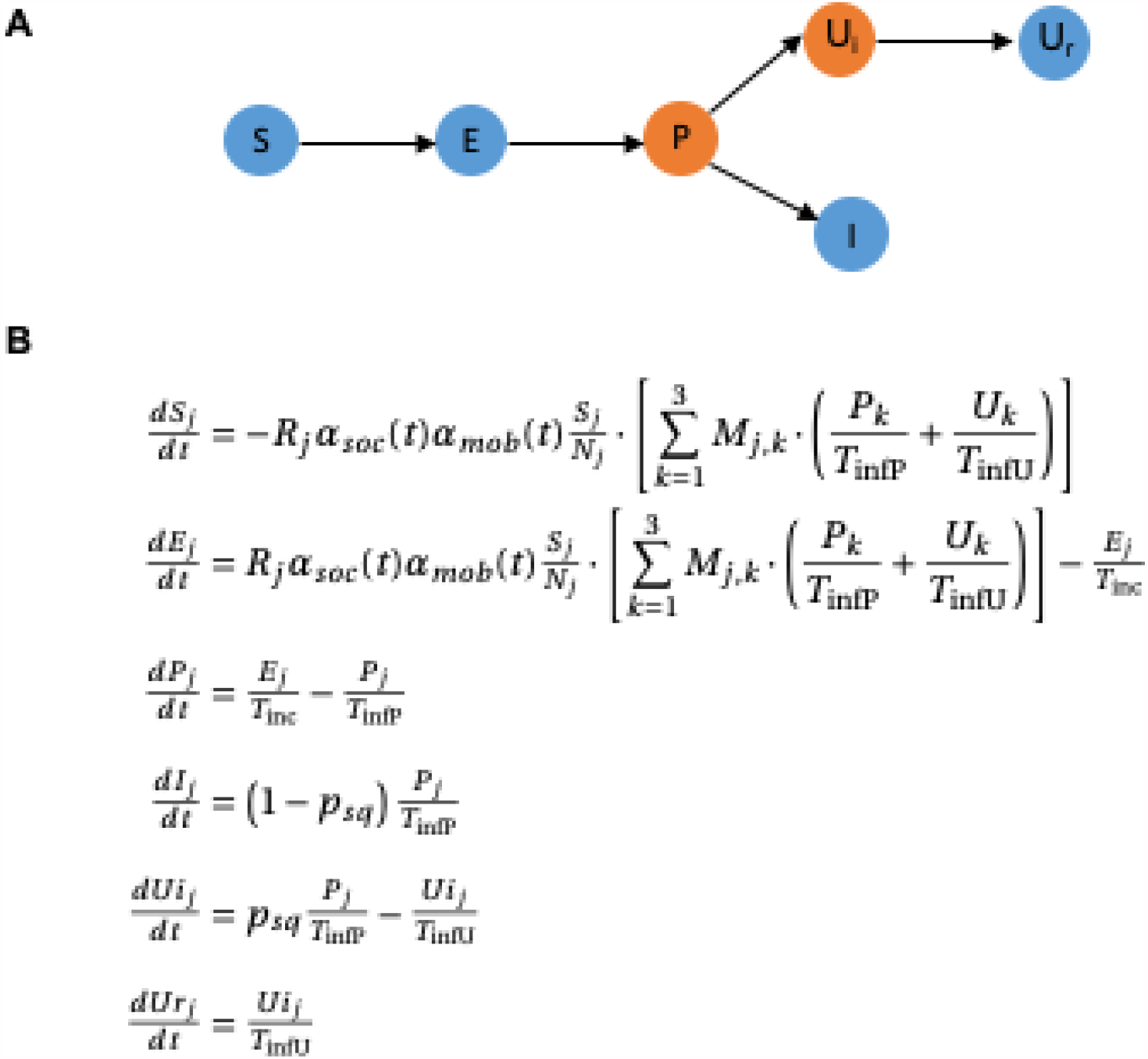
Overview of the SEIR model. A) Conceptual overview. We accounted for susceptibles (*S*), exposed (*E*, incubation time *T*_*inc*_), and pre-symptomatic yet infectious cases (*P*). After a presymptomatic time *T*_*in f P*_, cases were separated according to the estimated proportion of reported and sequenced cases, *p*_*sq*_, into either reported infectious (*I*), or unreported infectious (*U*_*i*_, reproductive number *R*). Since our data did not include information on recovered patients, a ‘recovered’ compartment was not included following *I*. It was assumed that reported cases remained isolated. The unreported compartment transitions to recovery (*U*_*r*_) after an infectious time *T*_*in f U*_. B) Relevant model equations to incorporate connectivity and exchange between the defined tertiles (index *j*). Cross contamination was included through the mobility matrix *M*_*jk*_ and relevant temporal variation of mobility and social interaction (weighting factors *α*_*mob*_(*t*) and *α*_*soc*_ (*t*)).

**Figure 2:**
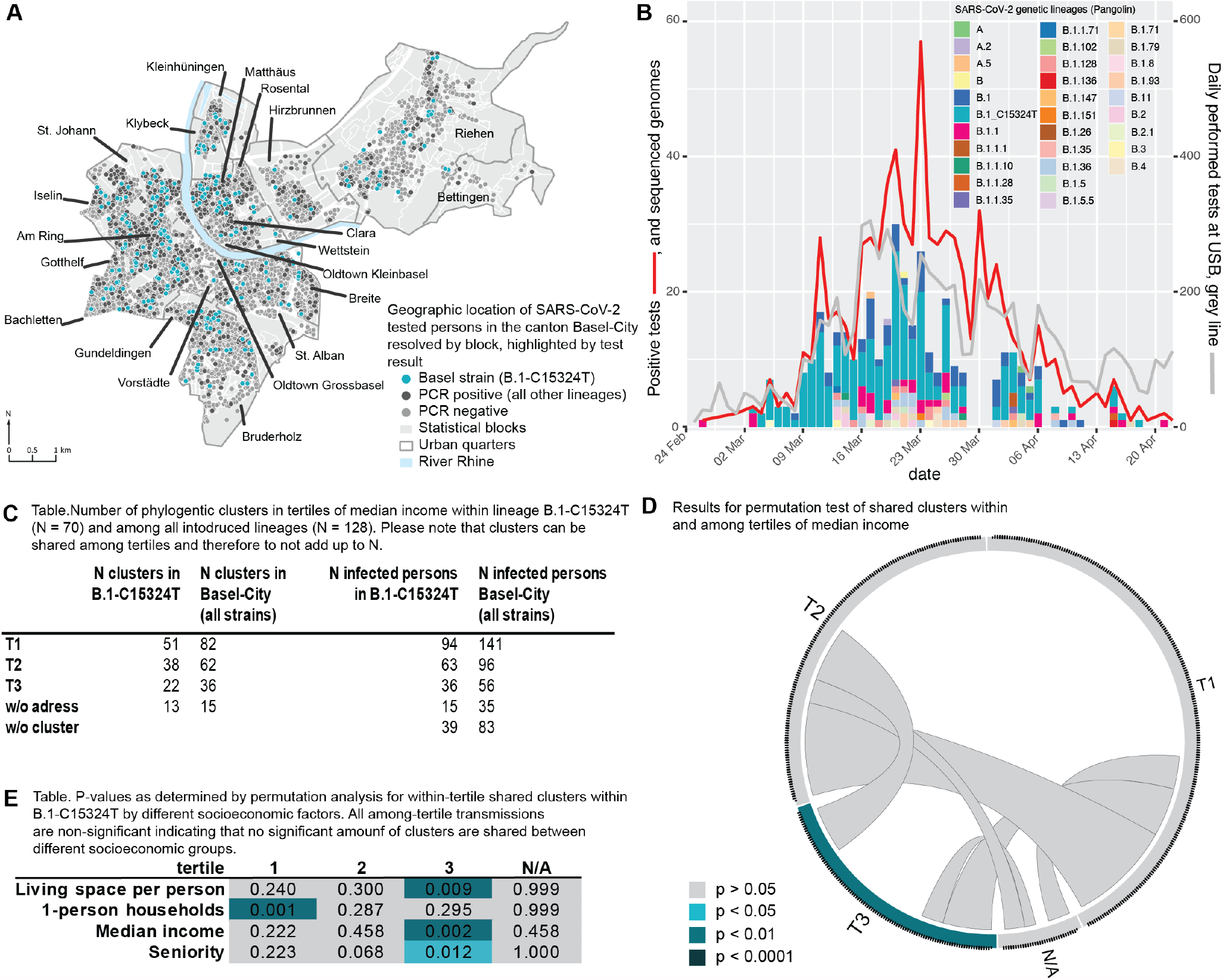
SARS-CoV-2 transmission in and among socioeconomic and demographic groups during the first COVID-19 wave in Basel-City. A) Spatial positive/negative case distribution throughout the city with the most dominant SARS-CoV-2 variant (B.1-C15324T), the focus of this study, highlighted in turquoise. B) Epidemiological curve for Basel-City and distribution of phylogenetic lineages (pangolin nomenclature) from 25^*th*^ of February to 22^*nd*^ of April 2020. C) Summary for inferred phylogenetic clusters within (i) all lineages and (ii) the major variant B.1-C15324T in tertiles of median income. High number of infected people within a tertile with a low number of clusters indicates presence of large transmission clusters whereas large number of clusters and low number of people infected within a tertile indicates random infections and cryptic transmission. D) Visualisation of a significance test for transmission within (indicated on circle edges) and among (intra circle connections) tertiles of median income. E) Results of a significance test for transmission between tertiles of different socioeconomic factors. T1: low, T2: intermediate, T3: high, N/A: no available data or censored for privacy reasons.

### Spatio-temporal variation of mobility and social interaction patterns

Figure 3A and Figures S3-S4 show Basel-City’s partition and the corresponding mobility graph. Importantly, the statistical blocks per tertile do not form a single, geographically connected, entity. We observe that mobility varies by transport modality and tertile (Figure 3A inset). For example, for low-median income (T1) the share of private motorized traffic and mobility is more pronounced than in the tertiles of higher median income (T2 and T3).

**Figure 3:**
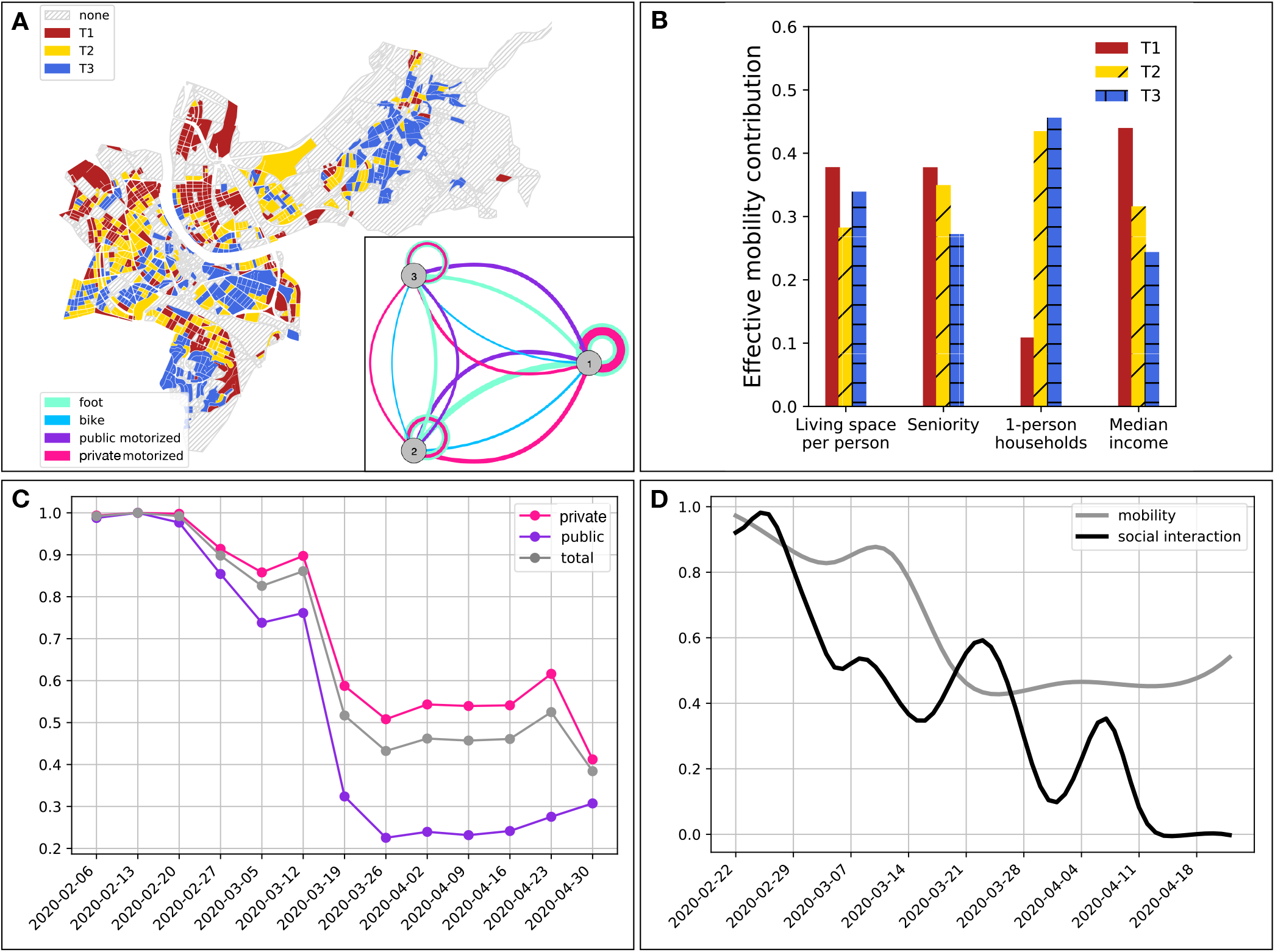
Spatio-temporal variation of mobility patterns within the Canton of Basel-City. A) Basel-City and its delineation with respect to statistical blocks colored according to the partition into tertiles T1, T2, and T3 of increasing median income. Inset: resulting mobility-graph, with nodes representing tertiles and edge-widths representing the strength of effective connectedness through mobility by means of various modes of transport, as computed from the traffic-model provided by the traffic department of the Canton of Basel-City. B) Relative mean contribution of mobility to a socioeconomic tertile’s effective reproductive number associated with the major variant B.1-C15324T. C) Normalized temporal development of private and public transport as well as their weighted sum during the first wave of the pandemic in Basel-City. D) Smoothed relative temporal development of social interaction and mobility contribution to the effective reproductive number associated with the major variant B.1-C15324T.

Figure 3B shows for each partition the summed edge weights of the mobility graph accounting for the mobility contribution to the final effective reproductive number. We observe that low and median income populations are more mobile than their wealthier counterparts. For living space per person or percentage of senior citizens, mobility was comparable between tertiles with a trend towards higher mobility within the younger population groups.

Dynamic changes in mobility were assessed by agglomerating normalized traffic counts for public and private transport modalities (Figure 3C). There was a clear drop in mobility for both public and private transport modes around the onset of the national lockdown date (12th March 2020). The decrease was more pronounced for public transport, resulting in a weighted average mobility drop of approximately 50% (Figure 3D). Figure 3D also shows the dynamic change in social interaction contribution to B.1-C15324T case numbers. Despite noticeable fluctuation, social interaction contribution decreased on average over time. This data also reflects variation in case reporting which affected the estimated effective reproductive number. Importantly, since the B.1-C15324T variant was eventually eradicated despite non-zero mobility, a final social interaction contribution of zero was expected.

### Spatio-temporal spread of the epidemic and its socioeconomic determinants

Unreported cases appeared to be a driving force of the transmission (88% for the sequenced B.1-C15324T variant). Figures 4A-C (and Figure S5) show the SEIR-model fit to data for each median income tertile. The corresponding dynamic change of the effective reproductive number (*R*_*eff*_) is given in Figures 4D-F. Independent of the underlying partition, the model provided adequate fits and we observed a drop in *R*_*eff*_ following the dynamic changes in mobility and social interaction. Importantly, there was a significant difference (achieved significance level below the Bonferoni corrected significance threshold of 5%) in *R*_*eff*_ between statistical blocks of the highest and lowest median income. For all socioeconomic partitions the obtained parameter distributions are summarized as histograms with median values and 95% confidence intervals in Figures 4G-Here, we found that blocks with higher living space per person, or higher median income had a significantly lower *R*_*eff*_ (< 1.7) compared to the maximum *R*_*eff*_ observed in the relevant partition. A partitioning based on the share of senior residents did not result in significant differences in *R*_*eff*_. Differences in *R*_*eff*_ are due to two factors: the effective mobility contribution (Figure 3B), and the modelled reproductive number (*R*, eq.(8)). In particular, the tertile with the highest share of median income (T3) showed less mobility compared to T2 and T3, emphasizing differences in *R*_*eff*_. By contrast, mobility in the T1 and T3 tertiles of living space per person were more similar (Figure 3B), yet differences in *R*_*eff*_ were significant, indicating that the transmission was not dominated by mobility alone.

**Figure 4:**
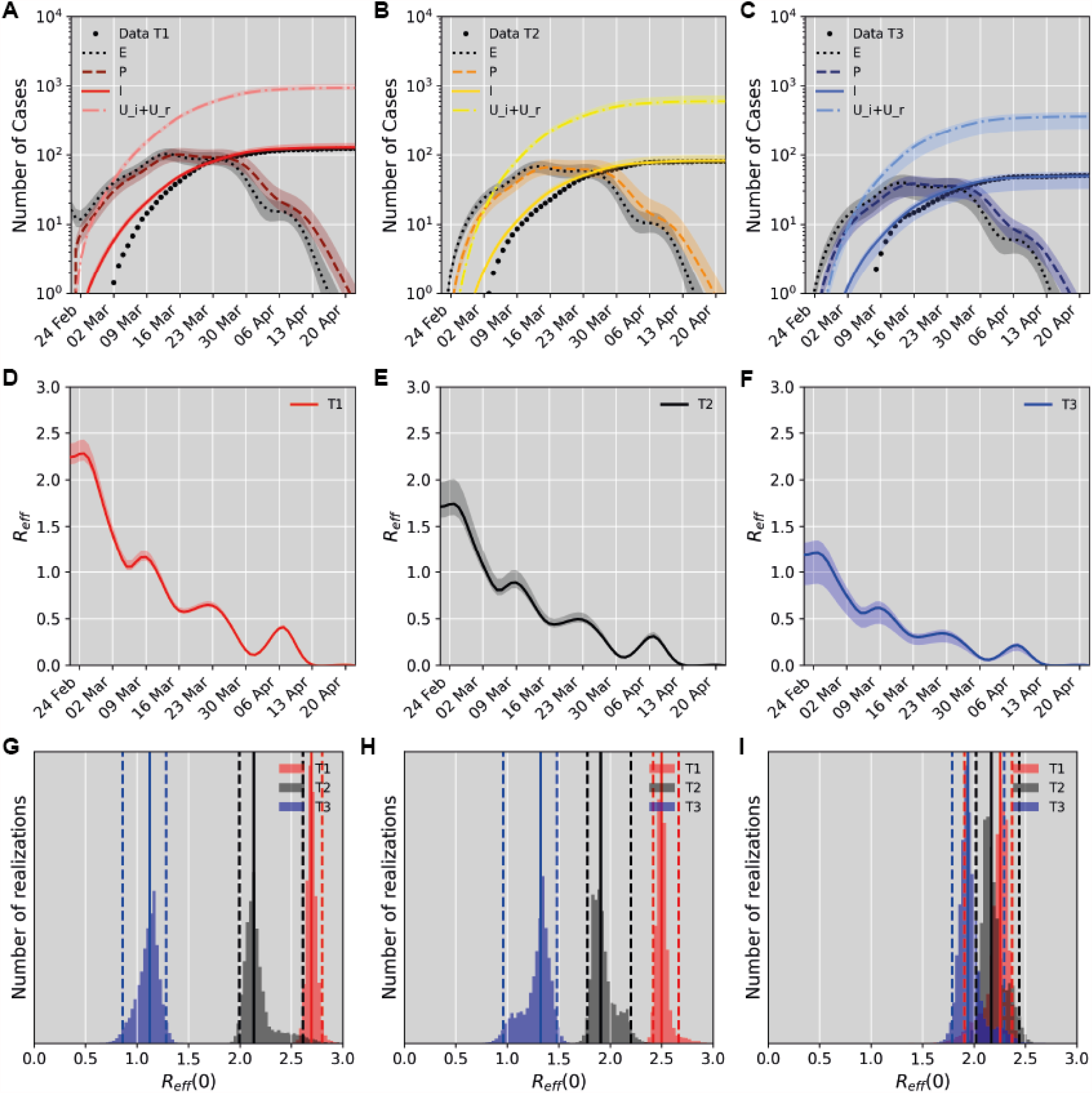
Model fit to the case number time-series. A-C) Fit results for a partition based on median income. Data points are shown together with model simulations (median (lines), and 95% confidence bounds (shaded)) for the different tertiles T1 (low, A), T2 (intermediate, B) and T3 (high median income, C). Model compartments *E* (exposed), *P* (presymptomatic), I (reported infectious), and *U*_*i*_+*U*_*R*_ (sum of the unreported infectious and recovered cases) are shown. D-F) The dynamic variation of the effective reproductive number for each of the tertiles shown in A-C. Median values (lines) are shown with 95% confidence bounds (shaded). G-I) Pre-lockdown reproductive number for each socioeconomic partition. Parameter distributions obtained using Markov Chain Monte Carlo analysis (histograms) are shown with median values (solid lines) and indicated 95% confidence bounds (dashed lines). Results are shown for partitions based on median income (G), living space per person (H), and share of senior residents (I).

### Impact of mobility changes and modelling of vaccination scenarios

We simulated the developments of the first wave of the epidemic under the assumption of different mobility scenarios and modelled two future vaccination strategies. Figure 5A displays the results for mobility scenarios as observed with up to 50% mobility reduction (scenario MO), 100% mobility (scenario M1), and no mobility (scenario M2). Peak case numbers (April 12^th^) would have been approximately three times higher in the case of no reduction in mobility (M1). Mobility reduction hence played a vital role for the containment of SARS-CoV-2.

**Figure 5:**
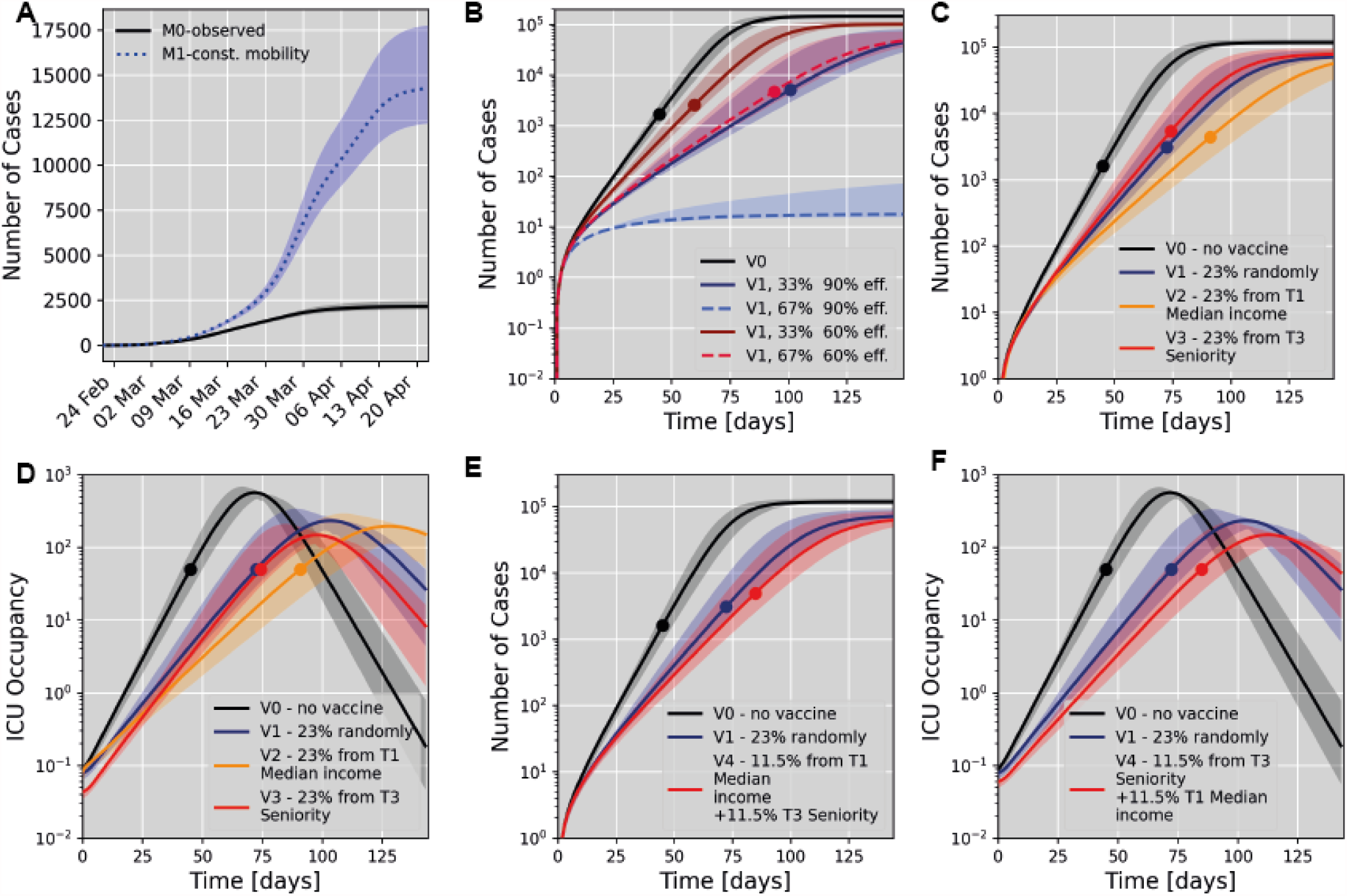
Scenario simulations for a partition based on median income. All subsets A,B,C and E show the total number of infected cases (i.e the sum of compartments *I, U*_*i*_, and *U*_*r*_. A) Influence of the mobility pattern on the total number of infected cases during the first wave (sum of reported and unreported cases) modelling either no change in mobility (no lockdown scenario, M1) or the observed scenario (MO) is shown. B) Simulation of simulated vaccination effects if a specific percentage of all citizens was randomly selected for vaccination at given efficacy (V1). We compare this to the scenario of no vaccine (V0). * Here vaccination of different population fractions at either 60% or 90% efficacy to prevent SARS-CoV-2 transmission were modelled. Median values (lines) and 95% confidence bounds (shaded) are shown. C) Simulation of future vaccination effects based on a partition according to median income. Scenario V2 models vaccination of 23% of all citizens selected from the tertile with the lowest median income (T1). Scenario V3 models vaccination of 23% of all citizens selected from the tertile with the highest share of senior residents (T3). In C-E we model 90% vaccine efficacy (transmission and severe COVID-19) and compare with scenarios V0 and V1. Dots indicate the time of reaching a 50% ICU occupancy. D) Temporal evolution of ICU occupancy for the scenarios modelled in C. E) Simulation of a mixed vaccination strategy giving equal priority to senior citizens and mobile population groups. F) Temporal evolution of ICU occupancy for the scenarios modelled in E).

Figure 5B shows the results for an outbreak scenario (denoted as V1) in which a specific fraction of the population (33% or 66%) received a vaccine that provided either 60% or 90% vaccine transmission reduction, with 90% severe COVID-19 case prevention. As expected, we observe that higher vaccine efficacy or higher population fraction vaccinated reduces the slope and plateau of the epidemic curve. Scenarios for less effective vaccines are shown in Figure S9. Effectiveness to prevent severe COVID-19 solely affects the rate of severe cases and hence ICU occupancy and the time point of reaching 50% ICU occupancy (Figure 5B vs. Figure S9E). It should be noted, that vaccination of the population at random is an artificial scenario, applied here only to demonstrate the impact of vaccination efficacy relative to the population fraction vaccinated. This scenario serves as a baseline comparison for two more realistic vaccination strategies given in Figure 5C. Here, as a proof of concept, vaccines that provide 90% transmission reduction, and 90% reduction of severe cases are delivered to 23% of the population are presented. For scenario V2, vaccination is prioritized in what we determined as determinants of SARS-CoV-2 transmission – individuals with low income, who have fewer options to socially distance (e.g. by working from home) and hence were more likely to be exposed to and/or transmit the virus (reflected by a higher *R*_*eff*_). With this strategy, the slope of the epidemic curve would be reduced compared to randomly vaccinating the same number of subjects from the whole population. Figure 5D describes the corresponding development of ICU occupancy for the scenarios modelled, revealing that scenario V2 leads to a delay of approximately 11 days to reach the 50% ICU capacity mark as compared to scenario V1. In scenario V3, resembling the approach by several countries, priority was given to the population group with the highest share of senior residents, which had lower mobility than the rest of the population (Figure 3B) but constitutes 60% of ICU cases. We observe that scenario V3 resulted in a marginally steeper epidemic curve (Figure 5C) and would yield 50% ICU capacity at a similar time as a random vaccination strategy (Figure 5D). However, the total number of cases at this time would be approximately double in scenario V3 compared to V1 (Figure 5C), whereas the overall peak ICU occupancy would be lowest in V3 (Figure 5D). These simulations suggest that - in case of vaccines reducing SARS-CoV-2 transmission - vaccination of population groups driving transmission are most effective in reducing the slope of the epidemic curve, whereas vaccination of high-risk groups reduces healthcare system burden in the long term. The presented effects strongly depend on the specific vaccine characteristics and population fraction vaccinated. In Figure 5E, F we finally show the potential effects of a mixed vaccination scenario giving equal priority to senior and highly mobile population groups as a representative example. This mixed mode provides a possible compromise with lower case numbers compared to V1, but also delayed and reduced ICU occupancy relative to V1.

## Discussion

This analysis evaluates complementary aspects of the spread of SARS-CoV-2 within a medium-sized European urban area, including local transmission analysed by phylogenetic tree inference and clustering, and the overall spread described by a compartmental SEIR-model enabling simulation of vaccination strategies. The main strength of this study lies in the high degree of diverse and detailed data included as well as the complementary models and analyses.

Patterns of SARS-CoV-2 transmission have previously been discussed from different angles either via network and transmission modelling ^24^, by statistical evaluation ^25^, or by phylogenetic clustering based on genomic sequencing data^26^. Independent of model choice, the importance of socioeconomic factors has been suggested previously^18,24^. However, analyses focused on metropolitan areas only may be biased towards their underlying socioeconomic and demographic characteristics making it important to also quantify SARS-CoV-2 transmission in other urban areas, such as in a city context, as well as in countries across all continents and stages of economic potential. Modelling studies provide the foundation of scenario predictions and estimation of effective reproductive numbers but require balancing the trade-off between detail described and the number of data points available. Accordingly, many published models rely on publicly available case numbers without being able to directly relate socioeconomic parameters and specific geographic locations per case, and/or are only performed for large metropolitan areas^19^. This has led to biases in evaluations and the neglect of a considerable population share living outside major cities (53%/41% in Europe/worldwide)^27,28^. In this study we chose Basel-City as a case study of a European urban area. Despite the strong economic status of Switzerland and an obligatory health insurance, implying potentially less extreme socioeconomic gradients than other countries, we demonstrate that socioeconomic background impacts the probability to acquire and transmit SARS-CoV-2. It would be expected that in cities containing more pronounced socioeconomic gradients, these disparities would emphasize transmission patterns of SARS-CoV-2 between and within socioeconomic groups.

The success of our analyses is based on the optimized choice of evaluation and model, high data-density and -quality, rather than large absolute case numbers. It is difficult to distinguish the spread of competing viral variants within the same population and to account for new introductions in a model since classical ODE or agent-based models are relying on the assumption of uninterrupted transmission chains. Sequencing information is essential to reliably inform on such transmission patterns. Yet given the cost of such analyses, WGS covering entire epidemic waves is often unfeasible. We included 81% of all reported cases in the study time frame and geographic area, which allowed us to restrict our analysis to a subset of 247 phylogenetically related cases consisting of a single SARS-CoV-2 variant. Given the ∼200k inhabitants of Basel-City, this implies one of the largest per-capita sequencing densities of reported studies to date. We deliberately choose a simple way to incorporate socioeconomic, demographic and mobility information into our compartmental model since more complex network approaches would be unfeasible for limited case numbers. The use of mobility and socioeconomic data in our models is unique since we include regularly collected data analysed by the statistical office of Basel-City, providing a high spatial and temporal resolution network of the inner city mobility patterns. In contrast to mobile phone data^16,19,24,29^ our data is not subjected to privacy legislation and is hence expected to be more readily available for other medium-sized urban areas around the world, making our analysis transferable. We do not hold information on the duration and specific location of individuals, but a continuum estimate of population mixing that aligns well with the concept of a compartmental ODE model. Mobility and the reduction thereof have been suggested as a proxy to evaluate the reduction of the spread of SARS-CoV-2 ^6,29,30^; however, there has also been a change in hygiene practices and social interaction behaviour. In our SEIR-model we separate these two contributions allowing for an easier translation of our model for scenario building. We further addressed unreported cases which were the driving force of infection outside the observed clusters. Our 77% estimate of unreported cases overall (not limited to B.1-C15324T) falls within the range of previous reports within Europe^8,31^. The SEIR-model evaluates general trends of transmission, such as effective reproductive numbers, and vaccine scenario building. To complement this, we employ phylogenetic analysis to identify transmission clusters. This comprehensive evaluation showed that socioeconomic brackets characterized by low median income and smaller living space per person, were associated with significantly larger effective reproductive numbers. In line with previous results^24^, we suggest that population groups from a weaker socioeconomic background are more mobile and at higher risk for SARS-CoV-2 infection/transmission originating from multiple sources via cryptic transmission. This aligns with the possibility that low socioeconomic status may relate to jobs requiring higher personal contact, and unavoidable mobility^32^, which has been shown to increase the risk of infection by 76%^33^. By contrast, phylogenetic clusters were predominantly discovered within higher socioeconomic, or more senior groups, implying a spread within the same social network. It is likely that those individuals are retired, or have had the ability to work from home, a pattern that has been observed also in other cities^34^. Effective contact tracing and testing strategies could be most efficient for these groups, which were not driving SARS-CoV-2 transmission.

These results should be accounted for during vaccine prioritization depending on the relevant vaccine characteristics and ICU capacity available. Our simulation framework provides flexibility to model various scenarios and vaccine efficacies but did not account for fatalities due to limited case numbers during the studied period in Basel-City. In case of a combined effect of vaccines to both prevent SARS-CoV-2 transmission and protect against COVID-19^35–37^, vaccination of individuals driving transmission in addition to the protection of high-risk groups would be ideal to arrive at a combined concept of protection from and containment of SARS-CoV-2. Vaccinating high-risk groups reduces the number of hospitalized and ICU patients in the short term, the spread of the pandemic would however, be more effectively contained by vaccinating the transmission drivers. By restricting vaccination to only risk groups, a larger fraction of the general population will be exposed to SARS-CoV-2 implying that contact and travel restrictions would remain necessary to contain transmission. Such measures come at great economic cost. Based on our results it would be recommended to follow a combined strategy to employ extensive testing where transmission chains are traceable, e.g. among less mobile population groups, and to combine this with a vaccination strategy aiming to prevent cryptic transmission.

It is important to clearly state additional assumptions and limitations of our approach to put it in context with previous studies. All data used in this analysis provide the highest level of detail achievable in the setting of an urban area, yet it they are far from individual level data or a large scale population level analysis. The estimate of the fraction of unreported cases and the assumption of it to be constant both over time and between population groups is a further simplification that was inevitable in light of the available data. It is assumed that testing rates may be biased towards socioeconomic levels. In contrast to other countries, COVID-19 testing was covered by the obligatory health insurance in Switzerland which may reduce, yet not fully prevent testing bias. It would be expected that the reported difference in transmission between socioeconomic groups may indeed be stronger than reported in this analysis. The choice of a continuum model excluded the possibility to account for stochastic superspreading events and we further did not account for continuous importations of cases. The presence of a significant superspreading event was indeed ruled out by our WGS analysis, whereas the restriction to a single variant predominantly reported in Basel-city limited the impact of potential case imports. Despite these limitations, we were able to obtain comparable results in terms of the impact of mobility and socioeconomic status as previously reported motivating the application of our approach for meaningful scenario building.

In conclusion, high-resolution city-level epidemiological studies are essential for understanding factors affecting pandemic transmission chains and thereby supporting tailored public health information campaigns and vaccination distribution strategies at the municipal level. We here provided an example of such an analysis within a representative medium-sized European city at the core of the Greater Basel area and part of the Upper Rhine Region Metropolitan Economy, which suggests that the findings and modelling approaches presented may be readily translated to other such areas.

## Materials and Methods

Detailed methods are given in the supplements.

### Included data

All analyses were based on PCR-positive (750/7073 tests) cases of residents of Basel-City between February 25^*th*^ and April 22^*nd*^ 2020, obtained from the University Hospital Basel, covering 81% of all reported cases in the relevant interval and location. All samples were subjected to WGS, 53% resulted in high quality genomes (i.e. 44% of all cases). Of these 247 (247/411, 60%) contained the monophyletic C15324T mutation in the B.1 lineage (B.1-C15324T) and were used for further analysis. Each case was linked to the patient’s place of residence anonymized to one of 1,078 statistical housing-blocks. For each of these housing-blocks, where privacy legislation permitted, Basel-City’s Cantonal Statistical Office provided socioeconomic indicators for the year of 2017 (most recent available). These included (i) living space (per capita in *m*^2^), (ii) share of 1-person private households, (iii) median income (CHF), and (iv) population seniority (percentage of citizens aged over 64 years). According to these indicators, blocks were allocated to one of three socioeconomic tertiles (T1:≤33rd percentile, T2: 33rd to 66th percentile, T3:>66th percentile, N/A: no available data or censored) where possible (e.g. Figure 3A). Generally, sparsely populated blocks displayed a maximum of three positive cases and had to be excluded from analysis. All following analyses with respect to socioeconomic factors were based on these partitions.

We determined SARS-CoV-2 antibody responses in 2,019 serum samples collected between 25^*th*^ of February and 22^*nd*^ of May 2020 to account for delayed seroconversion (see supplement for details). An estimated 1.9% (38/2,019) of the Basel-City population was infected with SARS-CoV-2, corresponding to 88% unreported cases for the sequenced B.1-C15324T variant (see supplement).

Finally, we included data on the number and age distribution of COVID-19 intensive-care unit (ICU) patients during the relevant period from University Hospital Basel, a tertiary hospital with a capacity of 44 ICU beds: 4.5% of reported SARS-CoV-2 positive cases were admitted to ICU and median length of ICU stay was 5.9 days (IQR, 1.5-12.9). 40% of these patients were younger than 64 years.

### Phylogenetic inference and cluster analysis

SARS-CoV-2 genomes were phylogenetically analysed in a global context as described previously^23^ (see supplements). Phylogenetic clusters were consolidated with epidemiological data (occupation in a health service job, resident of a care home, contact to positive cases, onset of symptoms, place of infection) to confirm the suitability of the divergence parameter chosen and then combined with ancillary geographic (quarter), and socioeconomic information as described above. We inferred statistical significance for clusters in respective tertiles.

### Mobility data

We employed the official traffic model provided by the traffic department of Basel-City ^38^ consisting of the 2016 average A-to-B traffic on a grid of ∼1400 counting zones for foot, bike, public motorized and private motorized transport. We computed the spatial variation of mobility within and between each of the socioeconomic partitions (see supplement) resulting in a unity-normalized three-by-three mobility matrix *M*_*jk*_ representing relative within-tertile/inter-tertile mobility on/off its diagonal. Additionally, weekly averages of pass-by traffic for combined foot-bike traffic, as well as private motorized traffic were obtained together with weekly public-transport passenger loads (from SBB Swiss Federal Railways and Basel-Verkehrsbetriebe). These were combined in a weighted sum according to the relevant transport mode contribution, normalized and smoothed with a uni-variate spline to obtain the final temporal mobility variation, *α*_*mob*_(*t*)).

### Dynamic changes in social interaction

SARS-CoV-2 transmission is contact-based. While the number of contacts is largely influenced by human mobility, the risk of a contact becoming a transmission event is further determined by the precautions taken by the individuals in contact (washing hands, wearing masks, distance keeping) resulting in an effective, time-dependent reproductive number *R*_*eff*_ (*t*). We derive the relevant time-dependence of *R*_*eff*_ (*t*) by applying a Kalman filter ^39,40^ to the piece-wise linearised time-series of daily confirmed B.1-C15324T cases. Assuming a multiplicative model, the time-dependence of transmission risk stemming from social interaction *α*_*soc*_ (*t*), is obtained by point-wise division of the time-dependence of *R*_*eff*_ (*t*) by *α*_*mob*_(*t*) (Figure 3D).

### SEIR-model

We used a compartmental two-arm susceptible-exposed-infected-recovered (SEIR) model ^19,41,42^ including sequenced and unsequencend/unreported cases that is outlined in Figure 1 using the following compartments: *S* (susceptibles), *E* (exposed, latency period *T*_*i*_*nc*), *P* (presymptomatic, infectious time *T*_*infP*_), *I* (reported infectious, isolated), *U*_*i*_ (unreported infectious, infectious time *T*_*infU*_), *U*_*r*_ (unreported recovered). The initial number of susceptibles was fixed to the relevant population. All other compartments were initialized as zeros, apart from a seed in *E* corresponding to the first reported cases. In summary, our model is based only on six free parameters: the reproductive number per tertile *R*_*j*_ (three parameters, range [0, 20]), the initial number of exposed in a single tertile (range [0, 20]), the infectious times *T*_*infP*_ (range [2., 12] days) and latency period *T*_*inc*_ (range [2., 12] days) ^43^. Since it was not possible to distinguish the fit for *T*_*infP*_ and *T*_*infU*_, we assumed a value of two days for the latter infectious time. The ODE system was implemented in python (version 3.8.) using the scipy functions *odeint* to iteratively solve the system of equations. Model fitting was performed on absolute (i.e. not cumulative) case numbers simultaneously for all partitions based on the least squares method using the *lmfit* library (version 1.0.2 ^44^). Posterior probability distributions of the fitted parameters were estimated using the Markov Chain Monte Carlo method implemented via the *emcee* algorithm ^45^. We report median values with 95% confidence intervals corresponding to the range of the 2.275th and 97.275th percentiles. We compare effective reproductive numbers corresponding to the normalization of *R* by the effective mobility contribution (Σ_*k*_ *M*_*jk*_):

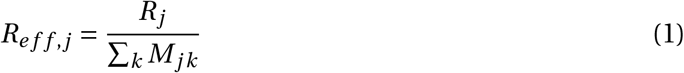

Significance levels of *R*_*eff*_ between tertiles are scored based on a comparison to 99 random partitions of the statistical blocks to a 5%, Bonferoni corrected significance threshold.

### Scenario simulation

The impact of mobility relative to social interaction was analysed by recalculating the predicted epidemic trajectory under the constraint of constant (*α*_*mob*_(*t*) = 1) mobility. This scenario was compared to the baseline of observed reduction in mobility. Vaccination scenarios were simulated for 70-90% effective vaccines to prevent COVID-19^46–48^, as well as 60% and 90% efficacies to prevent SARS-CoV-2 transmission. The effectively immunized fraction of the population was moved to the recovered compartment *U*_*r*_. We accounted for a change in social interaction behaviour following vaccination by assigning a mean social interaction score of the vaccinated and not-vaccinated population amongst the initial susceptibles (*α*_*soc,vacc*_ (*t*) = 0.75, *α*_*soc,novacc*_ (*t*) = 0.5). Full mobility was modelled (*α*_*mob*_(*t*) = 1) after a single exposed individual was introduced into each of the tertiles. All scenarios were compared to the no-vaccine case (V0): vaccination of a fixed population fraction at random (V1); vaccination of different population groups (V2: exclusively from T1 median income, V3: exclusively from T3 seniority, V4: 50% from T1 median income, 50% from T3 seniority). In addition to case numbers, the time to reach 50% of ICU capacity was determined as quantifiable endpoint indicating healthcare system burden. In case of not-random vaccination, we adjusted the relevant fraction of ICU cases based on the represented proportions of senior population fractions within all susceptibles.

## Data Availability

The SEIR-model code used for this submission will be available on https://github.com/BorgwardtLab/BaselEpi.git. Code that was used for phylogenetic inference and calculation of significance of clusters in specified groups is available at https://github.com/appliedmicrobiologyresearch. SARS-CoV-2 whole genomes from Basel-City are available at gisaid.com and at European Nucleotide Archive (ENA) under accession number PRJEB39887.

https://github.com/BorgwardtLab/BaselEpi

## Acknowledgements

We greatly appreciate the input and data received from Construction- and Traffic department Canton Basel-City, Baselland Transport AG, Basler Verkehrs-Betriebe, Autobus AG Liestal, SBB Federal Railways, Statistical Office of the Canton of Basel-City, and want to specifically thank Björn Lietzke, Lukas Mohler, and Madeleine Imhof (all Statistical Office of the Canton of Basel-City), from Construction- and Traffic Department of the Canton of Basel-City Michael Redle and Kathrin Grotrian, Matthias Hofmann (Basler Verkehrs-Betrieb), Roman Stingelin (Autobus AG), Nadine Ruch (SBB AG) and Stefan Burtschi (Baselland Transport AG) for their support. We thank Christine Kiessling, Magdalena Schneider, Elisabeth Schultheiss, Clarisse Straub, and Rosa-Maria Vesco (University Hospital Basel) for excellent technical assistance with next generation sequencing. Computations were performed at sciCORE (http://scicore.unibas.ch) scientific computing facility at the University of Basel. Data exchange was organized via the BioMedIT node between the University of Basel and ETH Zurich, Department of Biosystem Science and Engineering. We thank all authors, who have shared their genomic data on GISAID, especially the Stadler Lab from ETH Zurich for sharing Swiss sequences. A full table (csv) outlining the originating and submitting labs is included as a supplementary file. This study was supported by the Alfried Krupp Prize for Young University Teachers of the Alfried Krupp von Bohlen und Halbach-Stiftung (KB). We finally thank Dr. A. Jermy (Geminate Science Consulting) for his critical review of the manuscript.

## Author contributions

AE, HHH, KB devised the project. SCB and JK developed and performed the mathematical modelling. MSt performed and interpreted phylogenetic analyses. SCB, JK and MSt led the writing and revising of the report. AM provided the genome assembly pipeline. TCR and HSS prepared viral RNA for sequencing. MyB and RSS provided geographical expertise. RSS provided detailed background on the representative status of Basel-City. KKS collected clinical data. KR, DAT, AG, AKS, MSch analysed serology samples. DC and OD provided serology samples from Viollier AG. KL provided virological expertise. AB provided serology samples from the blood transfusion service. JB, STS and SF provided public health and epidemiological expertise. HP, MSi, CHN, RB, MB provided clinical expertise and valuable discussion on the results. NR, UH, JB provided epidemiological expertise. All author reviewed and edited the manuscript.

## Data Availability

The SEIR-model code used for this submission will be available on https://github.com/BorgwardtLab Code that was used for phylogenetic inference and calculation of significance of clusters in specified groups is available at https://github.com/appliedmicrobiologyresearch. SARS-CoV-2 whole genomes from Basel-City are available at gisaid.com and at European Nucleotide Archive (ENA) under accession number PRJEB39887.

## Ethical statement

Ethical approval was given by the local ethical committee *Ethik Kommission Nord-west und Zentralschweiz* (EKNZ No. 2020-00769, to be found at https://ongoingprojects.swissethics.ch) and the project was registered at clinicaltrial.gov under NCT04351503.

## Supplementary Material

### Research in context

#### Evidence before this study

We searched Google Scholar, PubMed, and medRxiv for articles with the keywords ‘SARS-CoV-2’, ‘transmission’, ‘phylogeny’, ‘city’, and ‘model’ as of February 2021 and evaluated the relevant abstracts and study details where appropriate. The identified studies predominantly evaluated SARS-CoV-2 transmission within large metropolises with only a few studies reporting on European cities, including analyses of London and Geneva. We noted, that studies (even outside Europe) generally differentiated between either phylogenetic analysis or a dynamic modelling approach and did not combine the two, despite the different aspects described. Moreover, the majority of studies evaluate publicly available data for modelling SARS-CoV-2 transmission and to estimate effective reproductive numbers. Such analyses naturally lack a direct relation of cases and relevant socioeconomic, or geographical information on a per case basis. To date, whole-genome-sequencing information has not been used to clearly identify inherently related cases for modelling transmission behaviour in any of the studies assessed. Finally, of the modelling studies reported, none provided a data-driven estimate of vaccine scenarios in a city, for which all model parameters were identified from the same source.

#### Added value of this study

In this comprehensive study we examine SARS-CoV-2 transmission patterns within a medium-sized European city. We benefit from a rich data set allowing us to combine phylogenetic clustering with compartmental modelling based exclusively on sequenced and inherently related cases that are mapped to a residential address, and hence coupled with socioeconomic and demographic information. This allows us to evaluate population groups driving SARS-CoV-2 transmission and to quantify relevant effective reproductive numbers. Moreover, we identify both traceable and cryptic transmission chains allowing us to suggest which measures (such as vaccination vs. effective contact tracing) would be most effective for each group.

#### Implications of all the available evidence

Depending on the specific characteristics of the vaccine, we estimate that vaccination of exclusively the senior population groups will reduce intensive-care-unit occupancy, but overall case number would more effectively be contained by prioritising highly mobile population groups from a socioeconomically weaker background (i.e. transmission drivers), even in the context of comparably shallow socioeconomic gradients within a wealthy European urban area. We identified predominantly clustered transmission amongst more senior or more affluent and less mobile population groups, which implies that extensive testing strategies could be used to more effectively prevent SARS-CoV-2 transmission among these groups. By contrast, mobile, low-income population groups were characterized by cryptic transmission. These are very important findings which could be considered in future vaccine prioritization designs for comparable urban areas.

### 1 Detailed Methods

#### PCR testing and whole-genome sequencing

PCR testing was available rapidly and frequent testing was established and supported by local guidelines by the end of February 2020, before the first case arrived^49^. SARS-CoV-2 testing was made available (i) via a walk-in test center in the city center affiliated to the University Hospital Basel, which allowed screening of legal-aged patients with mild and severe symptoms, (ii) via the University Children’s Hospital for minors on recommendation by the pediatricians, and (iii) via the obligatory screening of any incoming patients to the University Hospital Basel irrespective of symptoms. Testing in case of symptoms was covered by the Swiss mandatory health insurance scheme preventing sampling bias from affluent socioeconomic population groups. In total 7073 PCR tests from Basel-City residents were performed at the University Hospital Basel (UHB) (750 of which were SARS-CoV-2 positive) dating between 25^*th*^ of February and 22^*nd*^ of April, 2020. The total number of positive cases for Basel-City including also external testing sources for the same time range was 928, hence the cases registered at the UHB cover 80.8% of the total case burden^50^. The ratio of negative to positive PCR tests changed during the local epidemic with a median of 10.6% positive PCR tests (Figure 2B). We successfully sequenced SARS-CoV-2 whole genomes from 411 unique patients (54.8% of our cases, 44% of all cases). Of these, 247 (247/411, 60%) could be attributed to contain the C15324T mutation in the B.1 lineage (and therefore called B.1-C15324T) characteristic to the virus variant that originated in this tri-national area^23^.

#### Geographic mapping and socioeconomic stratification

Basel-City is divided into 21 urban quarters and had a population of 201,971 in 2020^51^ (Figure S6). For subsequent analyses, a total of 1,078 statistical (housing) blocks (a city block partitioned by e.g. streets, rivers) were identified within the city quarters. Each individual PCR test (N = 7073), irrespective of the result, was linked to the patient’s place of residence anonymized at the scale of statistical blocks in ArcMap 10.7 (by ESRI). To explore the statistical association between SARS-CoV-2 transmission and socioeconomic factors, we employed data provided by the Canton of Basel-City’s office for statistics for the year of 2017 (most recent data available), that specified the values for various socioeconomic indicators for each statistical block (except for those blocks where privacy legislation did not permit the sharing of such information). The indicators under study were (i) the living space (per capita in *m*^2^), (ii) the share of 1-person private households, (iii) the median income (CHF), and (iv) the population seniority (percentage of senior citizens aged over 64 years per block). According to these socioeconomic indicators, blocks were allocated to one of three socioeconomic city tertiles (T1:≤33rd percentile, T2: 33rd to 66th percentile T3:>66^th^ percentile) where possible (e.g. Figure 3A). In general, sparsely populated blocks displayed a maximum of three positive cases and had to be excluded from analysis. All following analyses with respect to socioeconomic factors were based on these city partitions.

#### Phylogenetic inference and cluster analysis

Whole-SARS-CoV-2-genomes from Basel-City patients were assembled using our custom analysis pipeline COVGAP^23^ (github.com/appliedmicrobiologyrese CoV-2/). Global sequences and metadata were downloaded from GISAID^52,53^ (as of October 22,2020; 155,278 consensus sequences). Sequences with more than 10 percent N’s (27,013) and with incomplete dates (43,466) were removed. 84,799 sequences remained which were joined with Basel genomes. Filtering for the period of interest until April 22^*nd*^ retained 39,913 genomes, of which 411 are from Basel-City residents dating from February 26^*th*^ (first case) to April 22^*nd*^, 2020.

To infer relatedness among the viral genomes and spread of SARS-CoV-2 in Basel-City, a time-calibrated phylogeny that was rooted to the first cases in Wuhan, China from December 2019, was inferred using a subset of the global genomes. For subsetting, we included 30 genomes per country and month, whereby all genomes from Basel-City were retained, totalling 3,495 genomes, using the nextstrain software v.2.0.0 (nextstrain.org) and augur v.8.0.0^3^ as described in detail in ^23^.

The resulting global phylogeny was used to infer phylogenetic clusters in Basel-City. First, polytomies, which are caused by identical genomes in the tree were resolved using ETE3 v.3.1.1^54^. Cluster Picker v.1.2.3 ^22^ was then used to identify clusters in the resolved tree (options 0, 0, 4e-4, 5). Identified clusters were consolidated with epidemiological data (occupation in a health service job, resident of a care home, contact to positive cases, onset of symptoms, place of infection) to confirm the suitability of the divergence parameter. Cluster Matcher v.1.2.4^22^ was then used to combine ancillary geographic (quarter), and socioeconomic or demographic information that were subdivided into tertiles on identified clusters.

To test whether related genomes in Basel-City cluster according to a) quarter, b) living space per person, c) share of 1-person households, d) median income, or e) seniority a custom python-script for a random permutation test was performed^55^ (github.com/appliedmicrobiologyresearch/Influenza-2016-2017). The results for clustering within and among urban quarter and tertiles in socioeconomic determinants were visualized using circos v.0.69^56^.

#### Serology

SARS-CoV-2 antibody responses were determined in a total of 2,019 serum samples collected from individuals between 25^*th*^ of February and 22^*nd*^ of May, 2020, to account for seroconversion. Serology information was used to estimate the fraction of unreported cases as follows: An estimated 1.88% (38/2,019) of the Basel-City population was infected with SARS-CoV-2. Of these 60% would be attributed to the B.1-C15324T variant, leading to a percentage of 88% of unreported/unsequenced cases to consider.

#### Mobility data

We employed the official traffic model provided by the traffic department of Basel-City ^38^. The latter consists of the 2016 average A-to-B traffic on a grid of ∼1400 counting zones for four transport modalities: foot, bike, public motorized transport and private motorized transport. We further obtained weekly averages of pass-by traffic for the same count zones over the period of the first wave of the pandemic for the categories of combined foot and bike traffic, as well as private motorized traffic. Additionally, weekly public-transport passenger loads were provided by both the Swiss Federal Rail Company and the local public transport services. From these datasets, we computed the spatio-temporal variation of mobility within the city as follows. Spatial variation was obtained by aggregating A-to-B traffic between the aforementioned counting zones, first to the statistical block level (by identifying the nearest housing block with respect to a zone’s centroid), and second to tertile level via a statistical-block’s association with a socioeconomic indicator tertile. This resulted in a there-by-three mobility matrix *M*_*jk*_ whose diagonal entries represent within-tertile mobility, while off-diagonal entries represent inter-tertile mobility. This matrix was normalized to one since only relative differences were relevant in our model. We hence obtained a direct link between mobility and socioeconomic/demographic information agglomerated based on the unifying scale of statistical blocks. Temporal variation was obtained by computing the weighted sum of the private transport time-series provided by the traffic department and the public transport time-series provided the Swiss Federal Rail Company and the local public transport services. This sum was then normalized and smoothed with a uni-variate spline resulting the final time-series for temporal mobility variation employed in our model (denoted as *α*_*mob*_(*t*)), see Figure 3D).

#### Dynamic changes in social interaction

SARS-CoV-2 transmission is contact-based. While the number of contacts potentially taking place within a day and a city is largely influenced by human mobility as estimated above, the risk of a contact becoming a transmission event is further determined by the precautions taken by the two individuals being in contact (such as washing hands, wearing masks, distance keeping). Both aspects together–mobility and risk-mitigating social behaviour within a (sub-)population–eventually result in an effective, time-dependent, reproductive number characterizing the virus’s transmission within that (sub-)population. Hence, there are three relevant time-series: changes in the overall effective reproductive number, in mobility, and in social behaviour. While the computation of the temporal variations in mobility was described above, the overall time-dependent effective reproductive number is obtained by applying a Kalman filter ^39,40^ to the daily case counts of individuals having newly contracted the B.1-C15324T variant of SARS-CoV-2 in all of Basel-City. To this end, we focus on the reduced dynamics as described by the *E, P*, and *U* compartments with constant times *T*_inc_, *T*_inf,P_ and *T*_inf,U_ (see Figure 1) obtained through a grid search. Changes in the number of susceptibles *S* is slow compared to the other compartments, which allows us to approximate that number as constant, and thereby linearize the dynamics–enabling the use of a Kalman filter in the first place. The measurement used to update the filter is *P* (*t*) as observed via the positive contribution *p*_sq_ · *P* (*t*)/*T*_inf,P_ to the daily increment of *U* (*t*). Assuming a multiplicative model, the time-dependence of residual transmission risk stemming from lack of precaution in social interaction (denoted as *α*_*soc*_ (*t*)), is obtained by point-wise division of the time-dependence of the effective reproductive number by the mobility time-series (depicted in Figure 3). Thus we are adhering to the logic that in the extreme case of zero mobility, no transmission can take place despite a finite risk of transmission rooted in a lack of precautions, while on the other hand in the case of zero risk of transmission due to perfect precautions, no transmission can take place despite non-zero mobility. Such logic dictates the choice of a multiplicative rather than additive model.

#### Fitting procedure and evaluation of the SEIR-model

In total 247 cases within the time period from the 25th of February until the 22nd of April were included in this analysis. For all data a seven day moving window average was taken to account for reporting bias on weekends. We fit absolute numbers of infected cases (*d I*). Due to the loss of single sequencing plate, missing numbers on the 29th, 30th and 31st of March were imputed by assuming a constant ratio of the B.1-C15324T variant amongst the sequenced samples. Simulations were initialized on the 22nd of February, the estimated date of the occurrence of the initial exposed cases^23^.

The ODE system was implemented in python (version 3.8.) using the scipy functions *odeint* to iteratively solve the system of equations. Data were fit using a least squares method implemented in the *lmfit* library (version 1.0.2 ^44^) with default parameters. Posterior probability distributions of the fitted parameters were estimated using the Markov Chain Monte Carlo method implemented via the *emcee* algorithm ^45^ of the *lmfit* library with default parameters. We report median values with 95% confidence intervals corresponding to the range of the 2.275th and 97.275th percentiles. The fit was performed simultaneously for all socioeconomic partitions to account for the shared parameters *T*_*in f P*_ (obtained 2.3(2.1,2.6) days), *T*_*inc*_ (obtained 2.4(2.3,3.4) days), and *E*_0_ (obtained 12.898.1,14.3)).

#### Scenario simulation

The impact of mobility relative to social interaction was analysed by recalculating the predicted epidemic trajectory under the constraint of constant intra-urban mobility (*α*_*mob*_(*t*) = 1, scenario M1) or fully restricted (*α*_*mob*_(*t*) = 0, scenario M2) mobility, corresponding to perfect isolation of the affected city areas. These scenarios were compared to the baseline of the actual reduction in mobility (scenario M0).

Vaccination scenarios were simulated as for both 90% and 70% effective vaccines to prevent COVID-19 resembling current vaccine candidate data ^46,47^, as well as a range of vaccine efficacies to prevent SARS-CoV-2 transmission (60%, 90%). This was achieved by moving the fraction of the vaccinated and not transmitting population from the susceptible to the recovered compartment *U*_*r*_ and calculating the spread of the pandemic with constant effective reproductive number and intra-city mobility. We accounted for a change in social interaction behaviour following vaccination by assigning a mean social interaction score of the vaccinated and not-vaccinated population amongst the initial susceptibles (*α*_*soc,vacc*_ (*t*) = 0.75, *α*_*soc,novacc*_ (*t*) = 0.5). Mobility was modelled as 100% (*α*_*mob*_(*t*) = 1). Two scenarios were investigated and compared to the no vaccine scenario (V0): i) vaccination of a fixed population fraction (one or two thirds) randomly throughout the population (scenario V1), ii) vaccination of the corresponding number of individuals from different socioeconomic groups (scenario V2 (exclusively from T1 median income), V3 (exclusively from T3 seniority), V4 (50% from T1 median income, 50% from T3 seniority)). In order gauge the benefit of a particular vaccination scenario, we calculated the time to reach 50% of intensive care unit (ICU) capacity. The University Hospital Basel has a total of 44 ICU beds. During the first wave, 4.5% of reported SARS-CoV-2 positive cases were admitted to ICU, and their median length of ICU stay was 5.9 days (IQR, 1.5-12.9). If considering additional unreported cases (captured by serological testing), the percentage of patients requiring ICU admission was 1%. Of all SARS-CoV-2 patients with ICU stay, 40% were younger than 64 years resulting in a probability of an under 64 year old infected case to be admitted to ICU of 0.5%. In case of vaccinated populations not at random, we adjust the relevant fraction of ICU cases based on the represented proportions of *geq* and < population fractions within all susceptibles.

#### Relevance of Basel-City in a European context - an overview

Basel-City has to be seen in its larger context: Basel-City is the core city of the trinational Greater Basel area and functional urban area (FUA), according to the official European statistics system. Basel-City is also part of a European cross-border region in the European Employment Services Network (EURES) (FigureS1) that promotes labor mobility across state borders and it is a main center in the context of “European Regions”. “European regions” are “designer regions” or spaces of cooperation within well defied action perimeters. The role of such forms of regional governance is to foster cross-border cooperation, regional competitiveness with collaborative development plans in interlinked, inter-locking metropolitan regions, which may be organized as “Eurodistricts” and are a mean in the EU-strategy to re-scale and decentralize development through regional institution building. Planning a the Eurodistrict level is regulated by state treaties and can act with a relatively high degree of autonomy from their national governments, mostly in the areas of spatial planning, traffic development and other aspects where links in development are missing.

Moreover, Basel and the Greater Basel area are also part of the Upper Rhine Region Metropolitan Economy, which by regional gross domestic product (GDP) is the eighth largest metropolitan economy in the EU. In summary, the case study of Basel-City is representative or transferable to other similar urban contexts because in the European harmonised statistical classifications (Eurostat and OECD classifications) as a metropolitan area, a functional urban area, and a part of a European cross-border region, it is representative for other urban areas classified similarly. This makes information obtained from analyses of Basel-City comparable and transferable to other such urban areas. This is explained in more detail below.

**Figure S1:**
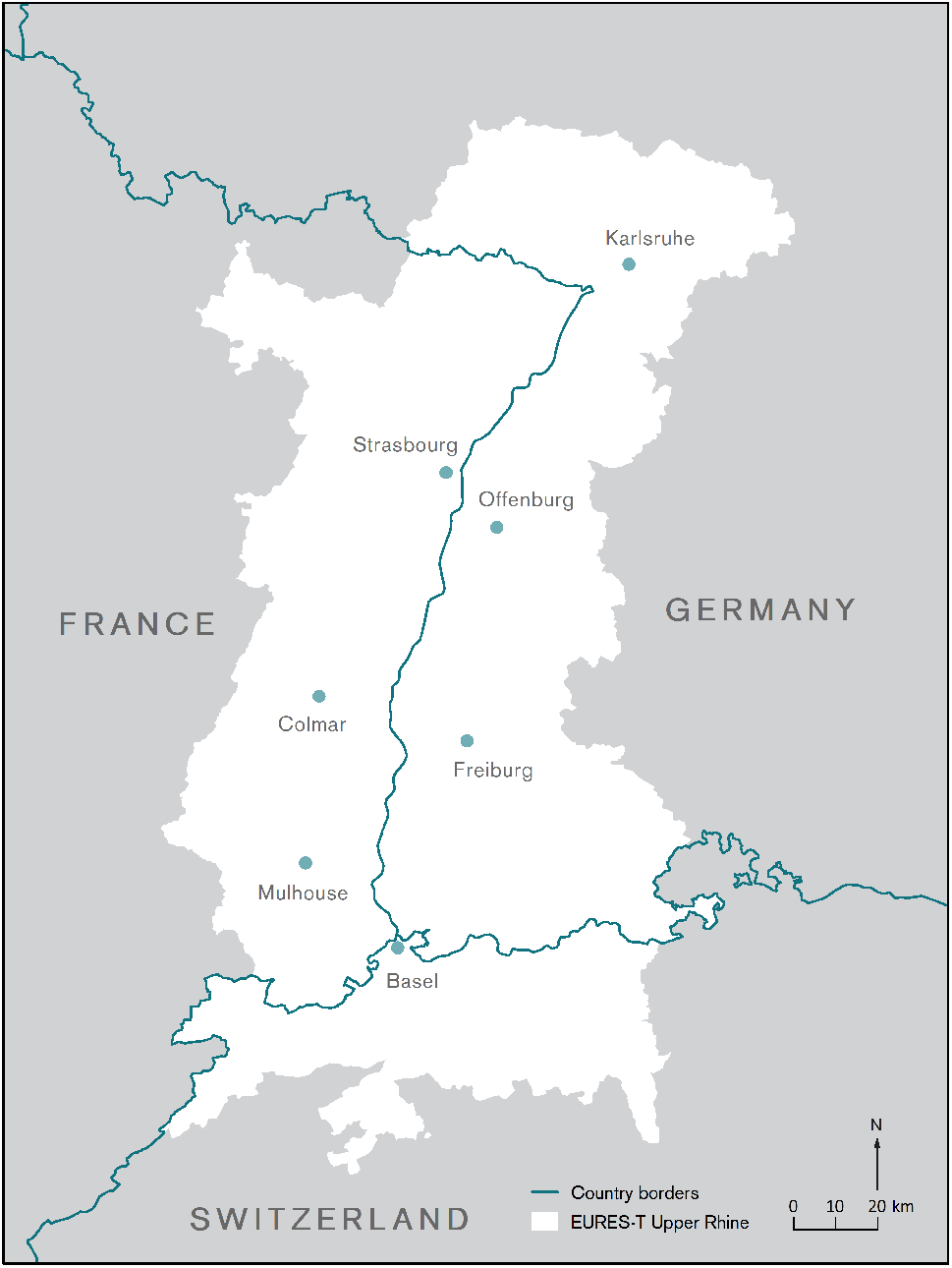
The EURES-T region Upper Rhine region for the purpose of promoting labor mobility across state borders. These comprise also the four “Eurodistricts” in the Upper Rhine “European region” for the purpose of decentralizing EU structural funds and promoting regional development. The Basel Trinational Eurodistrict is at the bottom.

#### Basel Metropolitan area

The Greater Basel area is a metropolitan area according the OECD/Eurostat definition, stretching into three countries with a population of around 830,000 in the Trinational Eurodistrict of Basel, an organization of municipalities and cities in the trinational surroundings of Basel and central cooperation body in the agglomeration of Basel ^57,58^. Metropolitan regions are NUTS 3 Eurostat statistical subdivisions according to the nomenclature des unités territoriales statistiques - a classification of territorial units for statistical analyses. The NUTS classification provides for a harmonised hierarchy of regions and Eurostat lists 541 such metropolitan areas ^59^.

Metropolitan areas are engines for growth and employment, the centers of competitiveness, and innovation, and contribute strongly to economic growth, social and political functions and are important for local, regional and international transport. Although only Basel-City, or rather, a small number of city quarters were examined here, Basel-City is a “hub”, i.e. the core of a larger metropolitan region with a commuter catchment area and on a scale that corresponds to the definitions of OECD and Eurostat (European Statistical Office) which also apply to Switzerland. As such, it is the contextualization of the city/urban quarter study within the larger urban/metropolitan/functional urban area, that make the study area comparable with other such metropolitan regions. Using Basel-City as a case study of an urban core or “core city” of its larger urban area / metropolitan area/ functional urban area makes the city, the study area and the results comparable or transferable to other urban areas/metropolitan areas/ functional urban areas and their core cities.

#### Basel as a hub of a Functional Urban Area

The Greater Basel area is also a functional urban area (FUA) in the official European statistics system. Functional urban areas consist of a densely inhabited city and a less densely populated commuting zone whose labor market is highly integrated with the city. FUAs extend beyond formal administrative boundaries. The OECD, in cooperation with the EU, has developed a harmonised definition of functional urban areas (FUAs). Being composed of a city (or core) and its commuting zone, FUAs encompass the economic and functional extent of cities based on daily people’s movements ^60^. The definition of FUA aims at providing a functional/economic definition of cities and their area of influence, by maximising international comparability and overcoming the limitation of using purely administrative approaches. At the same time, the concept of FUA, unlike other approaches, ensures a minimum link to the government level of the city or metropolitan area. The new harmonized OECD definition of cities, urban areas, functional urban areas and commuting zones allows for the first time a comparison within the European urban hierarchy. It identified 828 (greater) cities with an urban center of at least 50 000 inhabitants in the EU, and the “greater city level” greatly improved international comparability ^60^.

#### Basel-City as part of a European EURES-labor market and labor mobility region

Basel-City is also part of a European cross-border region within the European commission’s Strategy of Employment, Social Affairs and Inclusion European Employment Services-EURES cooperation network that promotes labor mobility in the EU and its partner countries in terms of assistance for recruitment and job placements, and providing information to cross order workers and employers on issues such as social security, insurance and taxation ^61^. EURES is based on technical standards and formats required for a uniform system to enable matching of job vacancies with job applications (Commission Implementing Decision (EU) 2017/1257 of 11 July 2017). EU internal border regions cover 40% of EU territory and are home to almost 2 million cross-border commuters. In 2018, more than 1.5 million people in the EU lived in one country and worked in another. In the trinational Basel urban area 60,000 persons commute on a daily basis across state borders, of which around 34,000 commute daily into Basel-City^62^. Just as mobility is an important factor in the economy, it may be a driving factor in the transmission of disease which is why the Basel study may be relevant for other cross-border regions in Europe.

#### Basel-City as part of a European cross-border region

The trinational Basel metropolitan area with Basel-City as its center has been the first to organize itself in private initiative as a “European cross-border region” in 1960 (Regio Basiliensis e.V.) for the purpose of advancing common interests and developments, enhancing cooperation between regions along state borders and between border regions throughout Europe. It has also served as a model for the institutionalised EU cross-border policy by means of INTERREG funding programs that aimed at promoting decentralized regional development through new forms of governance. Cross-border regions have flourished since the 1990 in particular because of their increasingly relevant role as implementation units for European regional policy in a context of multi-level governance^63^. Today, there are some larger 100 cross-border regions as outlined in Figure S2. Within the nested hierarchy of European “designer regions” for the purpose of fostering regional development some contain the aforementioned Eurodistricts, which are a rather new model of regional governance for promoting economic development in several contiguous metropolitan areas.

#### Perimeter of Unified Action of European regional governance

The Greater Basel area is furthermore part of the “Trinational Metropolitan Upper Rhine region” (TMUR). This refers to an innovative governance model for the four sub-areas Alsace, France, Northwest Switzerland, Southern Palatinate and Baden, Germany which together form an internationally strong business and knowledge location. The TMUR region refers to a perimeter of unified planning policy measures within a closely interlinked cross-border territory on issues of common interest. As a rather new form of territorial governance the TMUR region acts as umbrella over the existing metropolitan regions and trinational cooperations in the Upper Rhine and aims to strengthen their competitiveness within Europe and the world, and its public positions with respect to the political centers of Berlin, Paris, Bern, and Brussels. Collaborative efforts are focused on the areas of science, the economy, politics and civil society. By regional GDP and population size the TMUR can be an be seen as a major European metropolitan economy.

**Figure S2:**
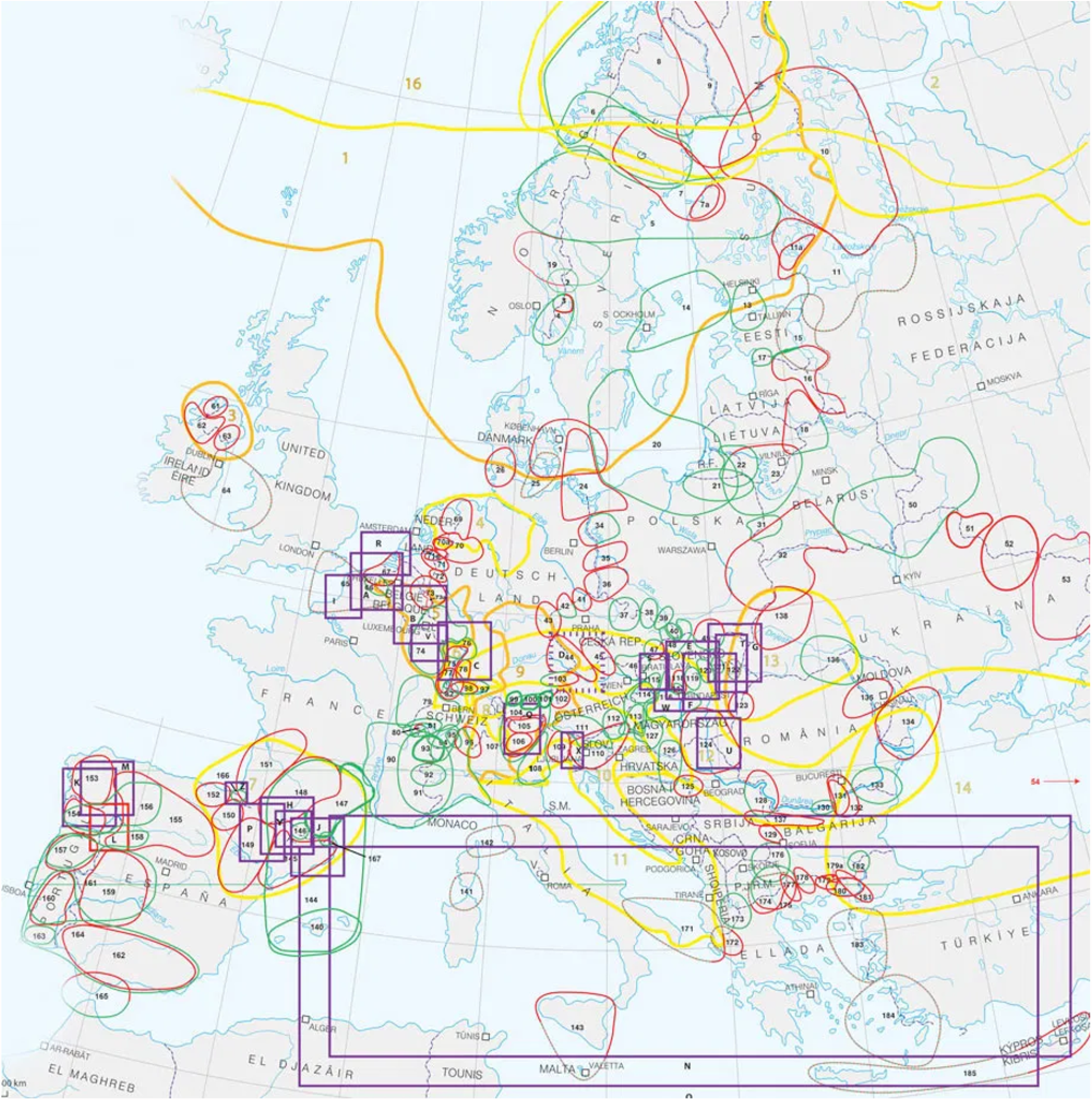
Association of European Border Regions (AEBR) and cross-border cooperation in Europe as shown in ^64^.

Metropolitan areas and metropolitan economies are engines and centers of growth and employment, and their many universities and knowledge institutions are drivers of innovation and international competitiveness. Large urban agglomerations typically combine economic, social and political functions and form important hubs for regional and international connection. Conceptually and methodically it is difficult to view this only in terms of administrative boundaries. The view on large metropolitan areas and regional governance models which encompass multitudes of cities and towns is applied for international comparisons, and in today’s global world the formation of larger metropolitan area governance forms which joins together urbanized areas even across administrative or state borders is the mechanism to achieve joint economic growth. The trinational metropolitan region Upper Rhine, in turn, is a major urban economy within Europe, ranking eighth in terms of regional GDP behind metropolitan areas outlined in Table S1.

**Table S1:** Selection of European metropolitan areas with GDP, population and GRP. Data obtained from ^65–68^

|  | GRP [bio. euros] | Population [mio.] | GRP per inhabitant |
| --- | --- | --- | --- |
| Greater London area | 946 | 19.53 | 48.443 |
| Paris (Ile de France) | 709 | 12.15 | 58.358 |
| Rhine-Ruhr area | 403 | 9.63 | 41.850 |
| Randstadt Netherlands | 397 | 8.15 | 48.659 |
| Milan (Lombardy) | 381 | 10.02 | 38.023 |
| Brussels-Antwerp | 264 | 5.67 | 46.656 |
| Barcelona (Catalonia) | 224 | 7.44 | 30.101 |
| **Trinational Upper** |  |  |  |
| **Rhine Metropolitan area** | **209** | **5.9** | **34.889** |
| Frankfurt (Darmstadt) | 200 | 3.95 | 50.666 |
| Copenhagen-Malmö | 180 | 3.29 | 54.827 |
| Munich | 177 | 6.12 | 79.690 |
| Berlin | 169 | 3.65 | 40.104 |

### Reasoning for the study design

Patterns of SARS-CoV-2 transmission have previously been discussed from different angles either via network and transmission modelling ^24^, by statistical evaluation ^25^, or by phylogenetic clustering based on genomic sequencing data^26^. Whereas modelling approaches can account for and simulate rich detail such as socioeconomic and demographic information, it is essential to balance the trade-off between detail described and the number of data points available. Moreover, a more diverse socioeconomic and demographic population structure may make it easier to detect general trends. In order to address these needs, many modelling studies rely on publicly available case numbers without being able to relate the cases to specific socioeconomic parameters and specific geographic locations, or have to perform analysis predominantly for large metropolis. For such scenarios, it is inherently difficult to distinguish the spread of competing viral variants within the same population and to account for new introductions in a model - classical ODE or agent-based models are relying on the assumption of uninterrupted transmission chains, information that whole-genome sequencing can provide. Yet given the cost of such analyses, whole genome sequencing covering entire epidemic waves is often infeasible. In this study we address the aforementioned limitations and choose a trade-off between the population detail studies in-light of limited case numbers for which we hold detailed information, including whole genome sequencing: we perform an analysis for a medium-sized, European city to account for the under-representation of these urban areas in previous modelling approaches. The case data used in this analysis was available for 80% of all reported cases in Basel-City and includes detailed, highly sensitive information on the infected individuals to enable tracing of transmission chains which are not publicly available. Sequencing was attempted for all samples and based on this data we restricted our analysis to inherently related cases of the B.1-C15324T variant. Although reducing the number of cases to <300, this implies that ODE based models are well suited to describe the dynamic development of these cases. We deliberately choose a continuum approach to incorporate socioeconomic, demographic and mobility information into this compartmental model since more complex network approaches would be infeasible for the final number of cases. This model enables us to evaluate general trends of transmission, such as effective reproductive numbers, and to use this information for vaccine scenario building.

In order to complement this more general analysis, we employ phylogenetic analysis to directly trace transmission clusters in detail. Phylogenetic analysis of positive cases provides rich information on relatedness of cases and hence supports modelling approaches by discerning introduction events and community spread. In this study we combine phylogenetic clustering with mathematical modelling of the SARS-CoV-2 transmission pattern. Our analysis is unique in the way that it is based on a large number of sequenced and phylogenetically related cases (411 sequenced out 750 positives, 247 that belong to a single genomic variant). As such, we however hold rich information for a comparably small, yet densely sampled cohort of cases, and are able to provide a complete picture of SARS-CoV-2 transmission in a medium-sized city by choosing two complementary analysis tools.

**Figure S3:**
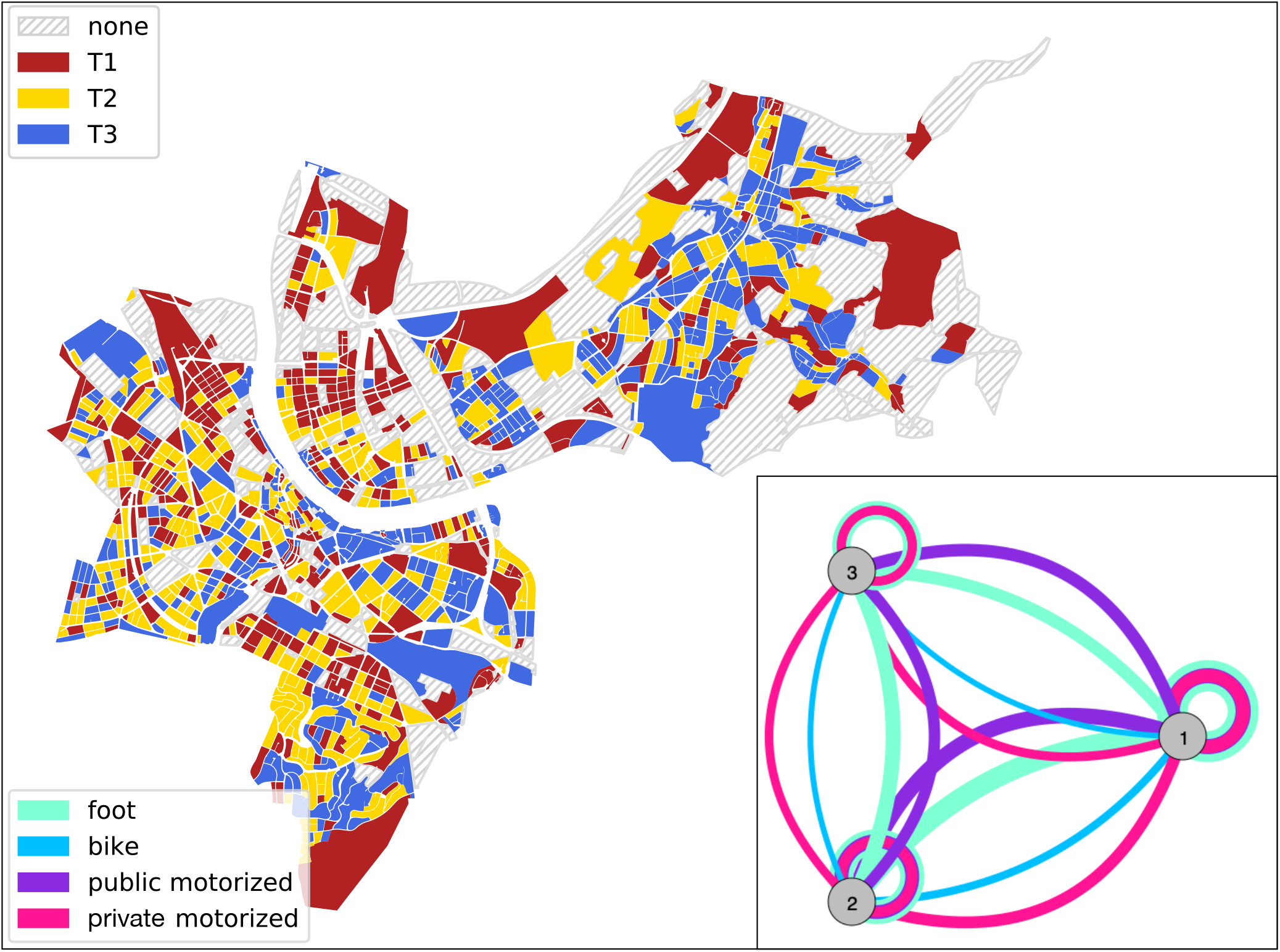
The Canton of Basel-City and its delineation with respect to statistical blocks colored according to the partition into tertiles T1, T2, and T3 of increasing fraction of residents aged older than 64 per block as provided by the canton’s office for statistics. Inset: resulting mobility-graph, with nodes representing tertiles and edges representing effective connectedness through mobility by means of various modes of transport (thicker/thinner edges indicating weaker/stronger connectedness), as computed from the traffic-model provided by the traffic department of the Canton of Basel-City.

**Figure S4:**
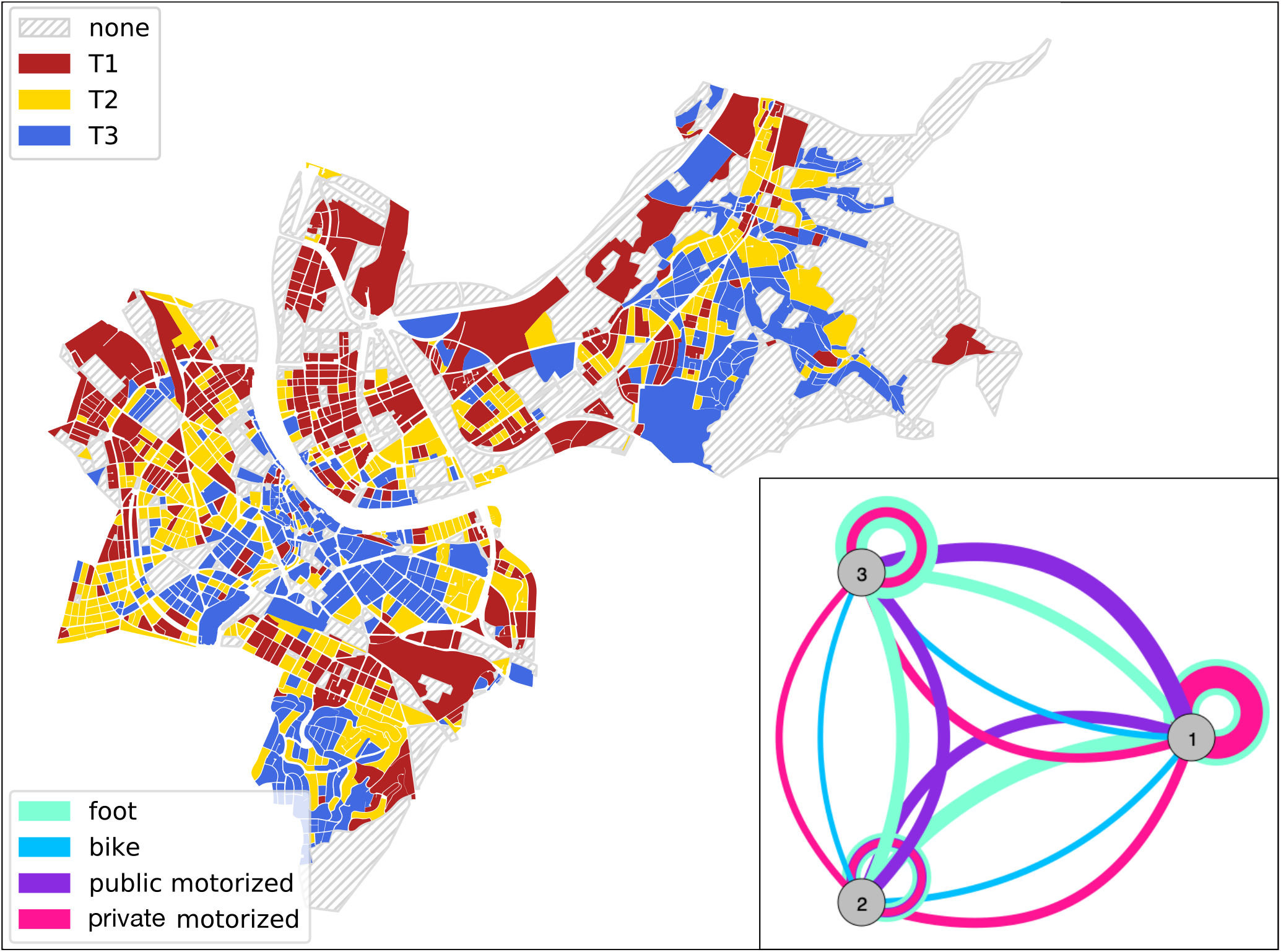
The Canton of Basel-City and its delineation with respect to statistical blocks colored according to the partition into tertiles T1, T2, and T3 of increasing living space per person as provided by the canton’s office for statistics. Inset: resulting mobility-graph, with nodes representing tertiles and edges representing effective connectedness through mobility by means of various modes of transport (thicker/thinner edges indicating weaker/stronger connectedness), as computed from the traffic-model provided by the traffic department of the Canton of Basel-City.

**Figure S5:**
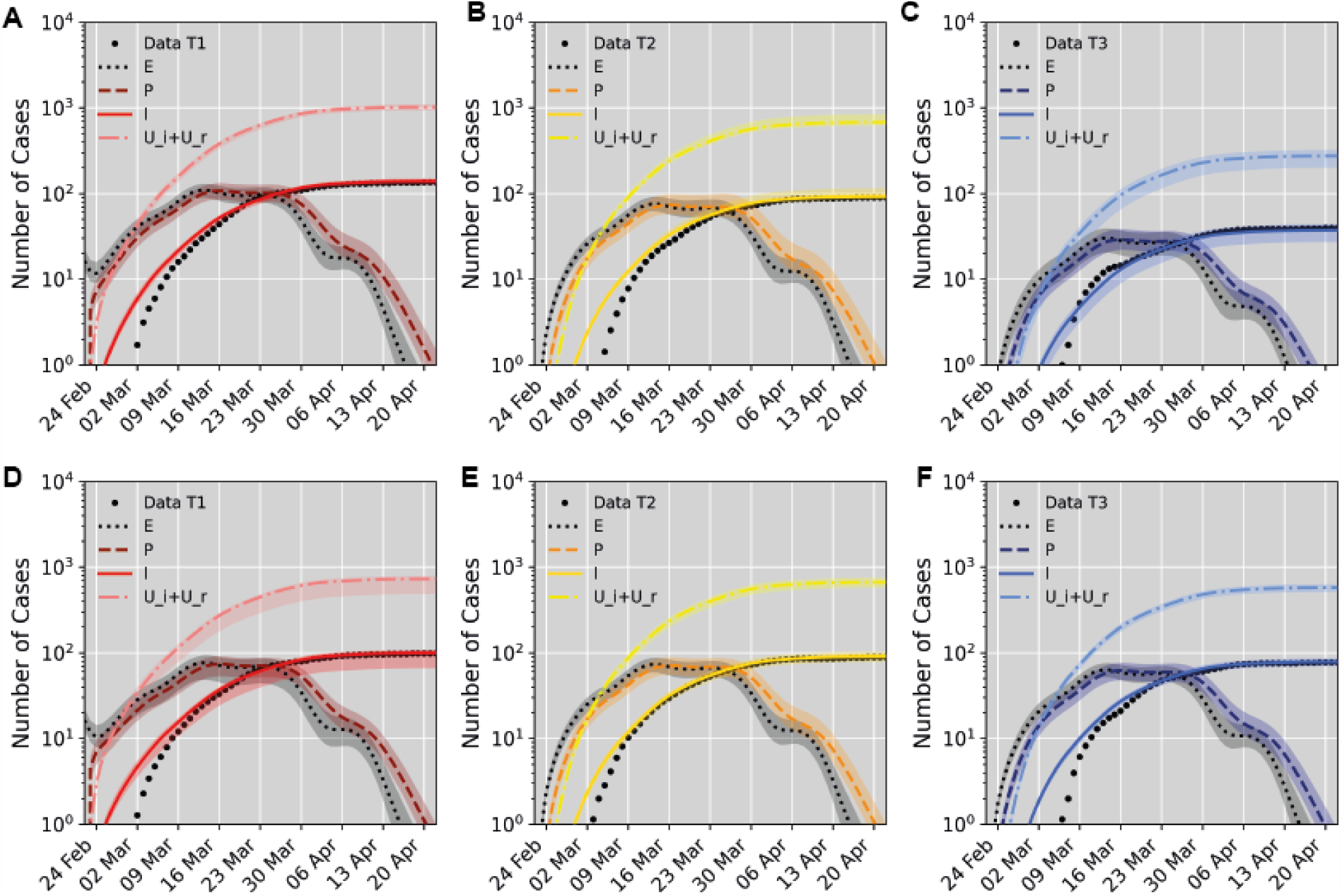
Model fit to data for partitions based on living space per person (A-C) and share of senior residents (D-F). Data points (dots) are shown together with model predictions for compartment I (solid lines, reported infected but isolated cases), the corresponding predictions for compartments *E* (exposed), *P* (presymptomatic) and *U*_*i*_+*U*_*r*_ (the sum of the unreported infectious and recovered individuals) shown as lines with 95% confidence bounds (shaded bands) based on 500 markov chains. Results are shown individually for each tertile T1(A, D), T2 (B, E) and T3 (C, F).

**Figure S6:**
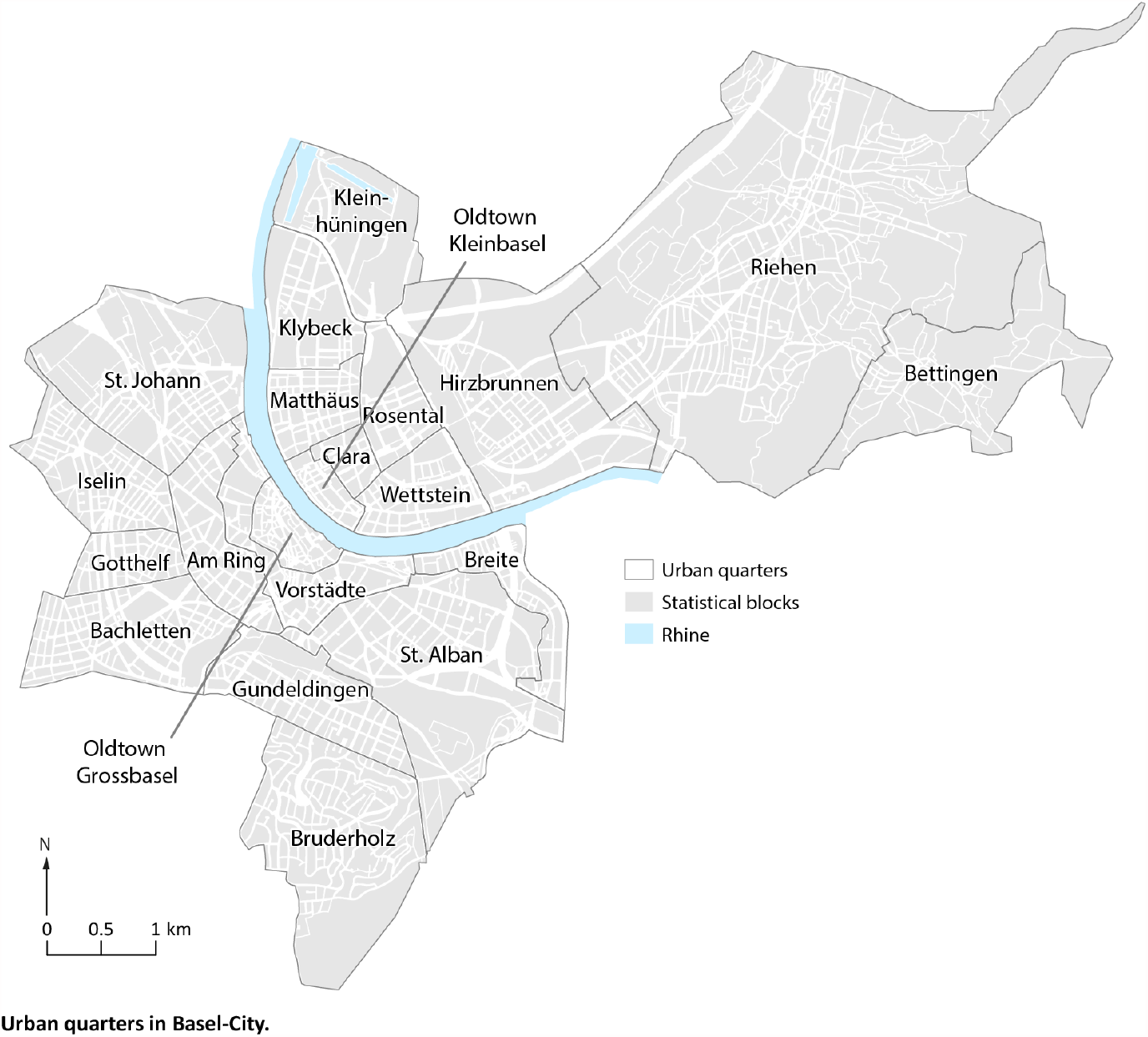
Urban quarters in Basel-City.

**Figure S7:**
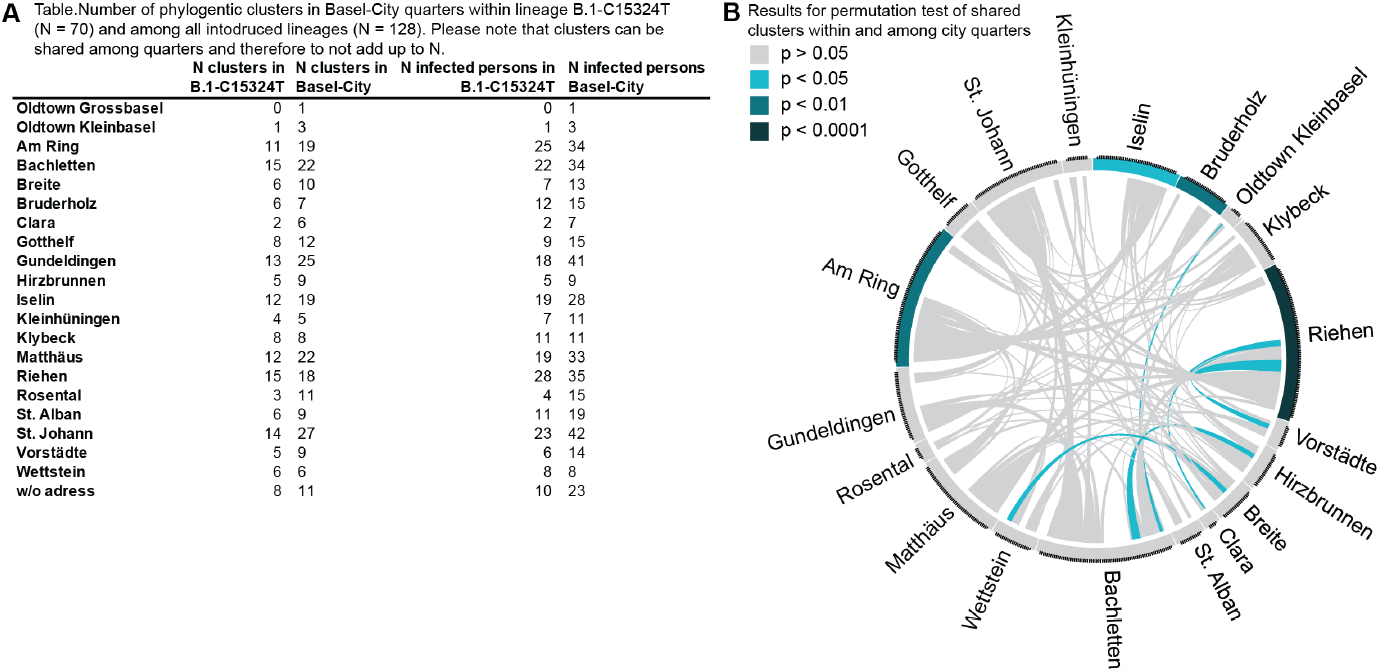
Phylogenetic clusters in quarters of Basel-City. A) Number of clusters within B.1-C15324T and within all viral variants in quarters of Basel-City. B) Visualisation of permutation analysis of shared phylogenetic clusters within and among urban quarters in Basel-City.

**Figure S8:**
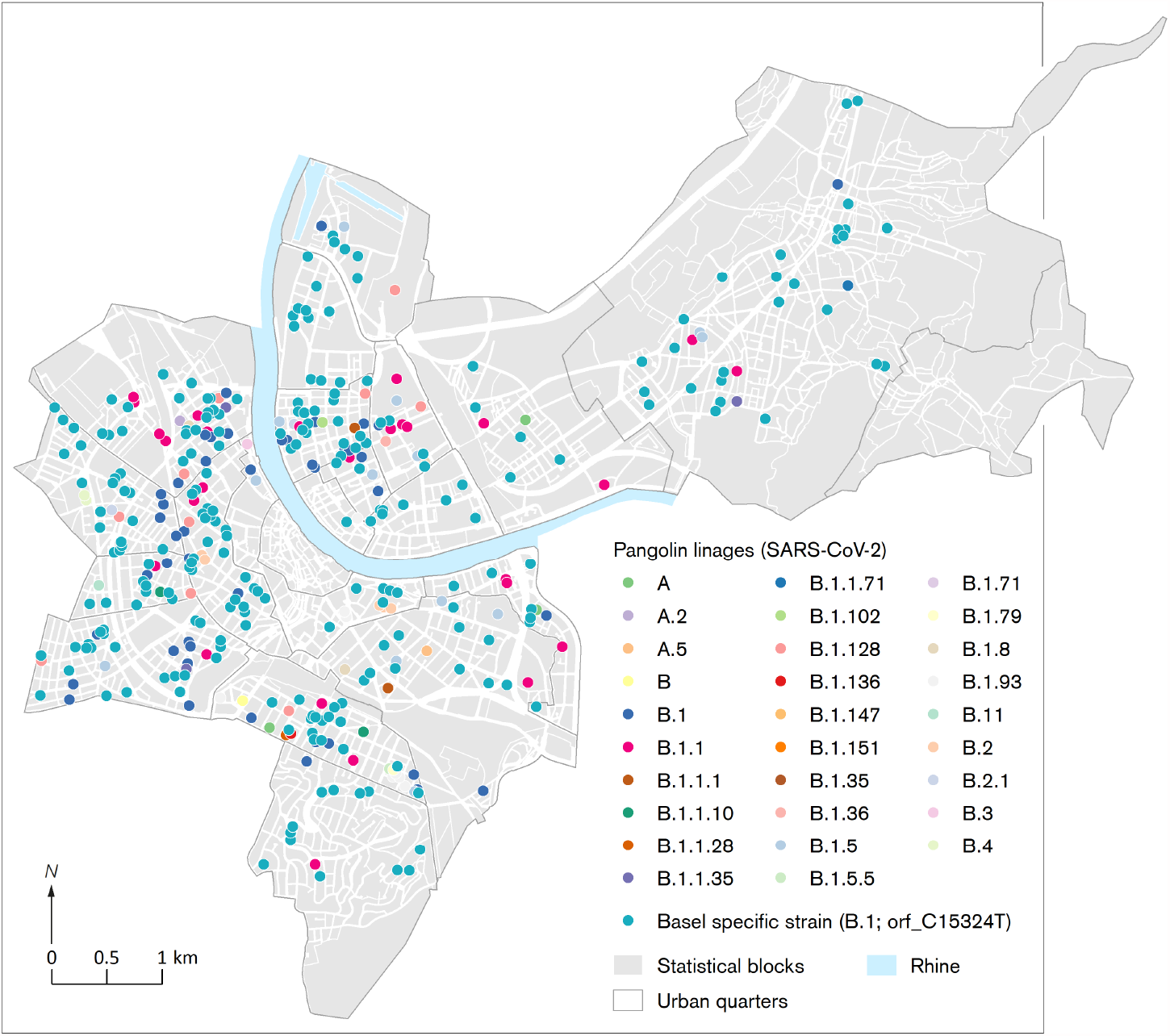
Lineage identity (pangolin) of PCR-confirmed COVID-19 cases from 26th of February until 22nd of April, 2020, in Basel-City with B.1-C15324T as dominant variant highlighted.

**Figure S9:**
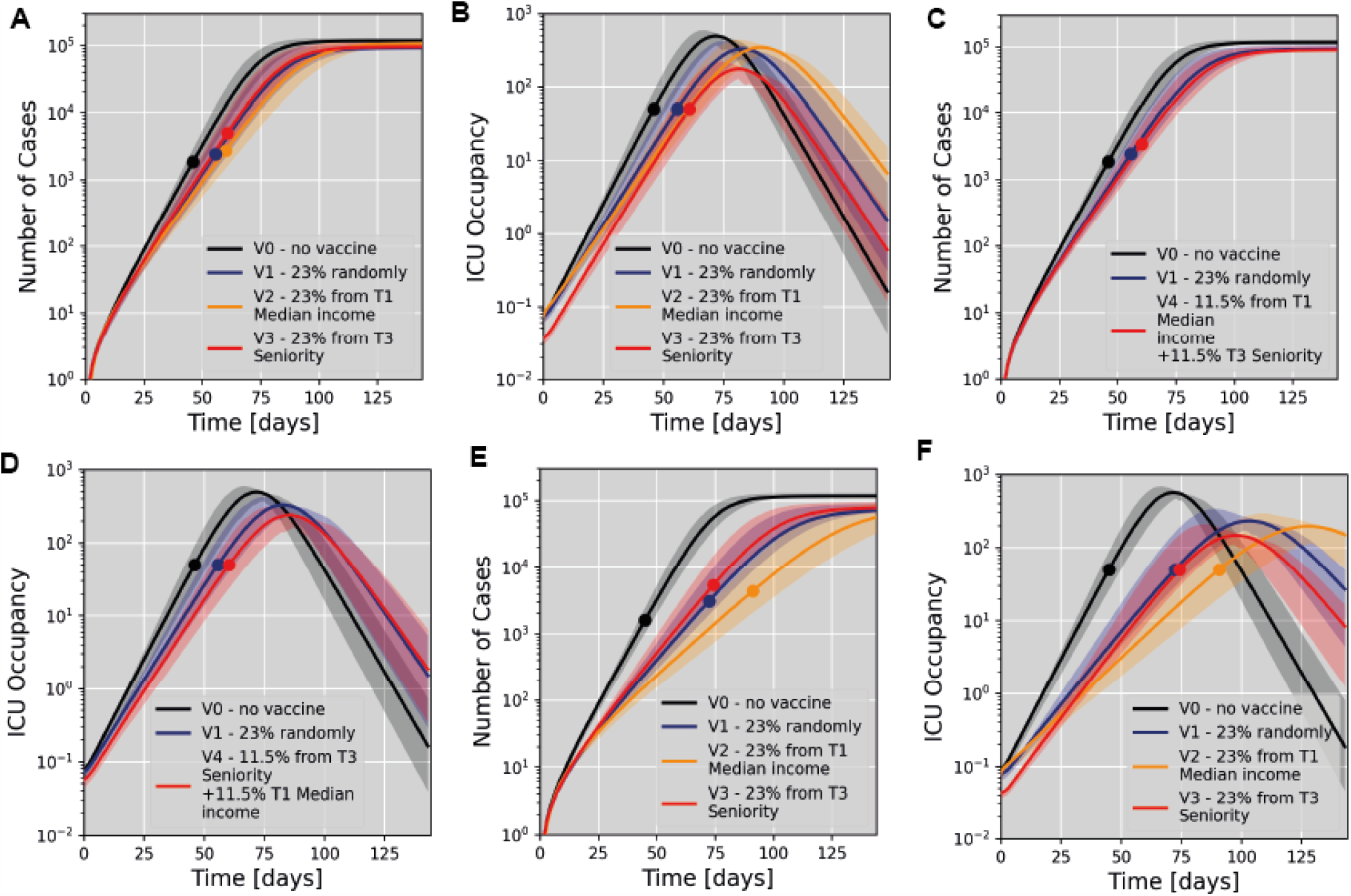
Modelling of vaccine scenarios assuming 60% (A-D) or 90% (E, F) vaccine efficacy to prevent SARS-CoV-2 transmission and 90% (A-D) or 70% (E, F) efficacy against severe COVID-19. We compare with scenarios V0 (no vaccination) and V1 (vaccination at random). Dots indicate the time of reaching a 50% ICU occupancy. A) Simulation of vaccination effects based on a partition according to median income. Scenario V2 models vaccination of 23% of all citizens selected from the tertile with the lowest median income (T1). Scenario V3 models vaccination of 23% of all citizens selected from the tertile with the highest share of senior residents (T3). B) Temporal evolution of ICU occupancy for the scenarios modelled in A). C) Simulation of a mixed vaccination strategy giving equal priority to senior citizens and mobile population groups. D) Temporal evolution of ICU occupancy for the scenarios modelled in C). E) Simulation of the same scenarios as in A) assuming 70% effective vaccination against severe COVID-19, and 90% vaccine efficacy to prevent SARS-CoV-2 transmission. F) Temporal evolution of ICU occupancy for the scenarios modelled in E).

